# Intra-individual reproducibility as essential determinant of clinical utility of smartphone-based neurological disability tests

**DOI:** 10.1101/2021.06.01.21258169

**Authors:** Komi S. Messan, Linh Pham, Thomas Harris, Yujin Kim, Vanessa Morgan, Peter Kosa, Bibiana Bielekova

## Abstract

Technological advances, lack of medical professionals, high cost of face-to face encounters and disasters such as COVID19 pandemic, fuel the telemedicine revolution. Numerous smartphone apps have been developed to measure neurological functions. However, their psychometric properties are seldom determined. Lacking such data, it is unclear which designs underlie’ eventual clinical utility of the smartphone tests.

We have developed the smartphone <u>Neu</u>rological <u>Fun</u>ction <u>T</u>ests <u>S</u>uite (NeuFun-TS) and are systematically evaluating their psychometric properties against the gold-standard of complete neurological examination digitalized into NeurEx^TM^ App. This paper examines the fifth, and thus far the most complex NeuFun-TS test, the “Spiral tracing”. We generated 40 features in the training cohort (22 healthy donors [HD] and 105 multiple sclerosis [MS] patients) and compared their intraclass correlation coefficient, fold-change between HD and MS and correlations with relevant clinical and imaging outcomes. We assembled the best features into machine-learning models and examined their performance in the independent validation cohort (56 MS patients).

We show that by aggregating multiple neurological functions, complex tests such as spiral tracing are susceptible to intra-individual variations, decreasing their reproducibility and thus, clinical utility. Simple tests, reproducibly measuring single function(s) that can be aggregated to increase sensitivity are preferable in app design.

## 1. Introduction

Expert neurological examination is an art that is slowly but surely disappearing. Skilled neurologist can reliably identify deficit in neurological function(s) and localize it to the specific part of the central (CNS) or peripheral nervous system (PNS). Expert examiner can also differentiate deficit that lacks anatomical substrate, by examining identical neurological function in different ways, noting inconsistencies, and motivating a patient to provide adequate effort. Such neurological examination takes between 30-60 minutes to perform and years to master. Because of examiner-dependency, the quantitative aspect of neurological examination, especially when performed by different raters, is less precise. Traditional neurological disability scales non-algorithmically aggregate semi-quantitative ratings of different neurological functions, usually selected by an individual (e.g., in Expanded Disability Status Scale; EDSS (Kurtzke, 1983)) or teams of experts (e.g., in Scripps Neurological Rating Scale; SNRS (Sipe et al., 1984)), into a single number. This is suboptimal for two reasons: 1. The features of the neurological examination aggregated to the disability scale are not data-driven and therefore may not be optimal and 2. Lack of defined algorithm causes errors during translation of the examination into a scale. These drawbacks are eliminated by data-driven scales (such as Combinatorial Weight-Adjusted Disability Scale; CombiWISE (P. Kosa et al., 2016)) and digital tools that allow convenient documentation of neurological examination in its entirety with automated, algorithmically codified computation of relevant disability scales (such as NeurEx^TM^ App (Peter Kosa et al., 2018)).

However, these solutions are useless when the lack of expert medical professionals, limited time for patient encounters or inability to examine patient in person due to pandemic, deprives patients of the benefit of this historically validated tool. Therefore, there is a strong movement to supplement neurological examination or, in some instances, to replace it, by patient-autonomous tests of neurological functions (both cognitive and physical) acquired via smartphones, tablets or web interphase (Balto, Kinnett-Hopkins, & Motl, 2016; Alexandra K. Boukhvalova et al., 2018; Bove et al., 2015; A. Creagh et al., 2020; Erasmus et al., 2001; Longstaff & Heath, 2006; Maillart et al., 2019; Midaglia et al., 2019; Pham et al., 2021; A. Vianello, L. Chittaro, S. Burigat, & R. Budai, 2017).

While some of these Apps are already marketed to patients, they often lack careful assessment of their psychometric properties against the gold standard of neurological examination and imaging or electrophysiological measures of CNS (or PNS) injury. In fact, even simple assessment of test-retest reproducibility may be missing.

Many of these Apps use tests adopted from standard neurological examination and modified to self-administered digital test. This is true for the Spiral tracing test, examined in this paper. Spiral tracing has been used in movement disorders to identify tremor and quantify its severity. Its digitalization offers automated identification and quantification of the tremor frequency and amplitude by Fourier transformation (Lin, Chen, Yang, & Chen, 2018). Furthermore, digitalization of the shape(s) tracing allows other quantitative measurements of speed and precision of the tracing (by finger or stylus) which may reflect neurological (dys)functions.

We have reviewed previous studies of digitalized spiral/object tracing (A. Creagh et al., 2020; Erasmus et al., 2001; Feys et al., 2007; Longstaff & Heath, 2006) to derive comprehensive set of digital features (40 total) and determined their psychometric properties (i.e., reproducibility, ability to differentiate MS patients from HD and correlation with relevant features of neurological examination, disability scales and CNS tissue destruction visible on brain MRI) in the training and independent validation cohorts of MS patients. Our data indicate that smartphone modification of spiral tracing has inferior clinical utility compared to simpler smartphone tests in NeuFun-TS, such as finger tapping (Tanigawa et al., 2017), balloon popping (Alexandra K. Boukhvalova et al., 2018) and level test (Boukhvalova et al., 2019), likely because of its complexity and resulting poor reproducibility when modified to smartphone self-administration.

## 2. Materials and Method

### 2.1 Participants

The NeurEx data were collected from participants enrolled in the Natural History protocol: Comprehensive Multimodal Analysis of Neuroimmunological Diseases in the Central Nervous System (Clinicaltrials.gov identifier NCT00794352). The study was approved by National Institute of Allergy and Infectious Diseases (NIAID) scientific review and by the National Institutes of Health (NIH) Institutional Review Board. All methods were performed in accordance with the relevant guidelines and regulations. All study participants gave informed consent. HD were recruited in two ways: 1) full participants in the Natural History protocol that underwent comprehensive neurological/imaging evaluation and 2) participants in a substudy of the Natural History protocol to obtain normative data for smartphone apps (without neurological/imaging evaluation). Two different groups are comprised within the MS datasets: a cohort that is tested in a clinic approximately every 6 months (non-granular testing sub-cohorts) and those that had the smartphone at home and did the test more than 5 times during a period of two years (granular testing sub-cohorts). Prior to all analyses, the MS datasets were separated into a 2/3 training and 1/3 test set weighted by one of the clinical features (average 9HPT; see Table 2 in Section 2.3). A summary of the demographic information is provided in Table 1.

**Table 1.**
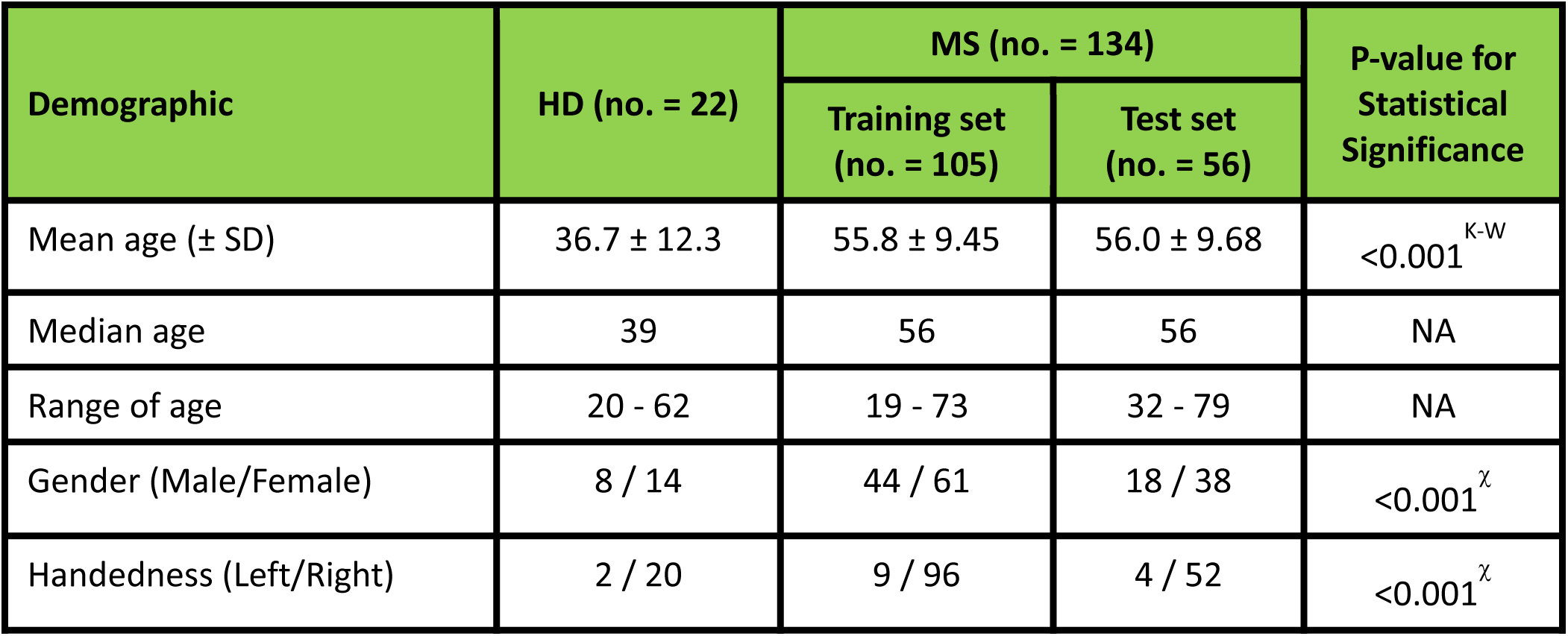
Demographics and characteristics for Healthy Donors (HD) and Multiple Sclerosis Patients over the 2 years study period. SD indicates standard deviation from the mean. K-W indicates Kruskal-Wallis non-parametric test for the mean comparison of age across HD, MS-Training set, and MS-Test set. χ denotes the Chi-Square test of independence of the differences between categorical groups (i.e., gender or handedness in HD, MS-Training set, and MS-Test set). A subset of MS patients in the Training set are also in the Test set but with different test dates. NA denotes not applicable. MS denotes Multiple Sclerosis.

**Table 2.** List of clinical disability scales used in the study. ✔ marks indicates clinical disability scale (i.e., clinical features) that are statistically significant different between HD and MS at the Benjamini-Hochberg (BH) adjusted p-value of 0.001. Disability scale that are not statistically significant between HD and MS are marked with dashed symbols (-). 18 out of the 22 clinical features were statistically significant between HD and MS. HD and MS denote Healthy Donors and Multiple Sclerosis respectively.

| Label | Feature Description | Statistically Significant |
| --- | --- | --- |
| C1 | 9HPT Average | ✓ |
| C2 | Non-dominant hand 9HPT | ✓ |
| C3 | Dominant hand 9HPT | ✓ |
| C4 | Expanded Disability Status Scale (EDSS; 0-10; ordinal) | ✓ |
| C5 | CombiWISE (0-100; continuous) | ✓ |
| C6 | NeurEx (0-1349; continuous) | ✓ |
| C7 | EDSS Visual Function Score | ✓ |
| C8 | NeurEx Vision Score | - |
| C9 | EDSS Pyramidal Functions Score | ✓ |
| C10 | NeurEx Pyramidal/Motor Functions Non-dominant hand | ✓ |
| C11 | NeurEx Pyramidal/Motor Functions Dominant hand | ✓ |
| C12 | EDSS Cerebellar Functions Score | ✓ |
| C13 | NeurEx Cerebellar Functions Non-dominant hand | ✓ |
| C14 | NeurEx Cerebellar Functions Dominant hand | ✓ |
| C15 | NeurEx Vibration and Proprioception Non-dominant hand | ✓ |
| C16 | NeurEx Vibration and Proprioception Dominant hand | ✓ |
| MRI1 | Brainstem atrophy | - |
| MRI2 | Medulla/Upper C-spine atrophy | - |
| MRI3 | Cerebellum atrophy | - |
| MRI4 | Lesion Load Brainstem | ✓ |
| MRI5 | Lesion Load Medulla | ✓ |
| MRI6 | Lesion Load Cerebellum | ✓ |

### 2.2 Test Design and Data collection

The Spiral test was written in Java and Kotlin using the Android Studio integrated development environment. The test is distributed as an Android Package (APK) over email, or directly installed to the device over USB, and updates are sent out over the air.

The testing devices Google Pixel XL and Google Pixel 2 XL, running Android (Android Version 11), with the intent of keeping them up to date. Results are uploaded to Firebase Firestore, a commercial cloud database, with alphanumeric identifiers to avoid Personally Identifiable Information. Spirals are generated using physical dimensions and rendered using the individual device’s screen characteristics and configuration, to ensure that spirals with the same parameters look the same across all devices.

Spiral tracing test consisted of tracing with a finger of each hand an orange spiral shown on the screen of the smartphone at three difficulty levels: Level 1 (simplest) consisted of thickest spiral of shortest length, while level 3 (most difficult) consisted of thinnest spiral of longest length (Figure 1). Each participant was instructed to trace the spiral as accurately and fast as possible. A total of four test trials were conducted by the subjects at each of the test dates for each difficulty level. Two of the drawings are done clockwise from and to the center of the spiral by the dominant hand and similarly, two drawings are done by the non-dominant hand counterclockwise (i.e. from and to the center of the spiral). During the experiment, a raw sensor data was collected from the smartphone touchscreen as an x- and y-screen coordinates with a corresponding timestamp in milliseconds and an estimated pressure of the tap based on the surface area of the finger on the touchscreen.

**Figure 1.**
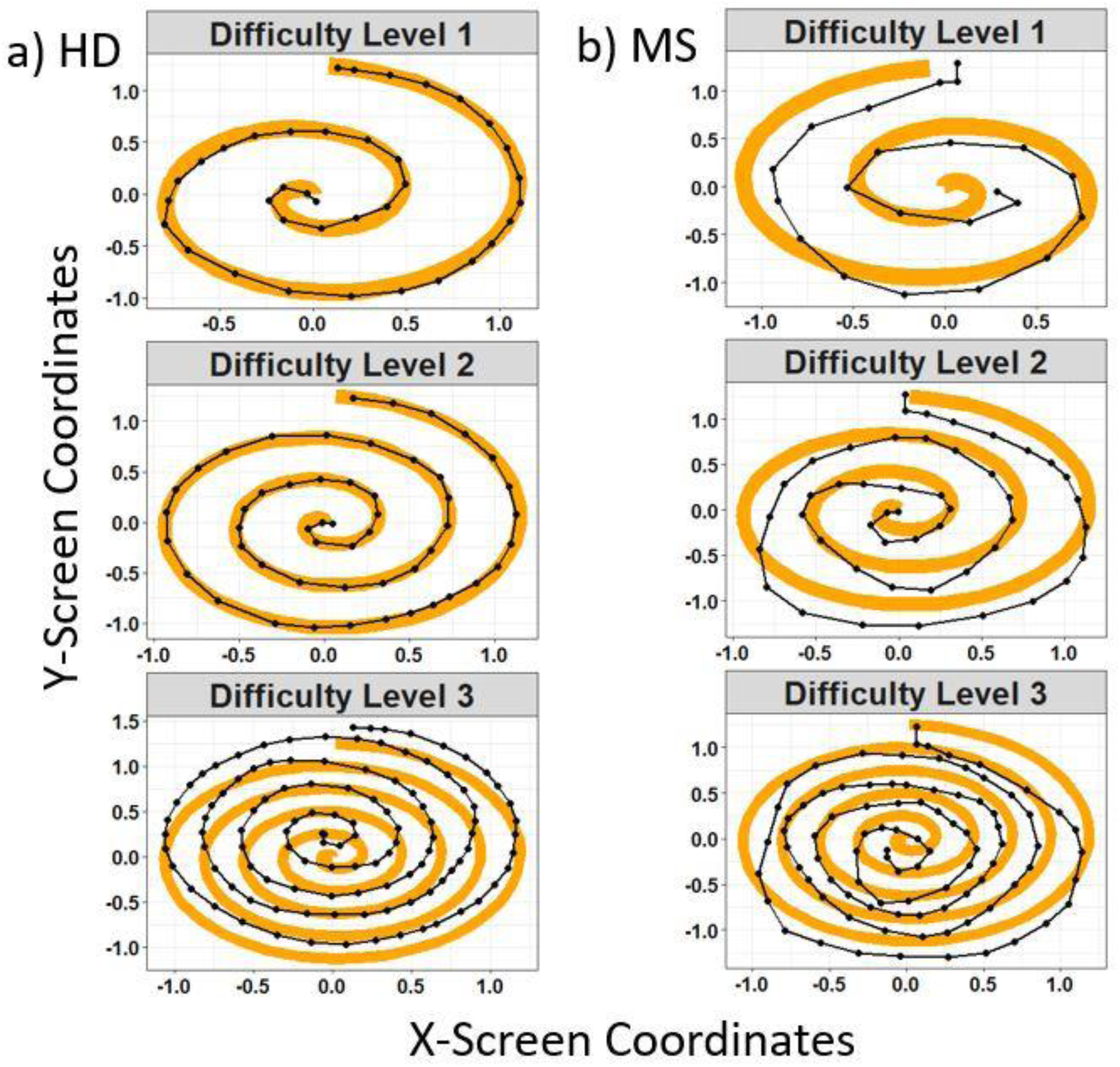
Representation of the spiral tests performed on the smartphone Apps by healthy donors (a) and MS patients (b). The orange and black spirals represent respectively the reference shape and the patient’s drawn shape. MS denotes Multiple Sclerosis

### 2.3 Clinical assessments of motor symptoms

The complete neurological examination, lasting 30-60 minutes and performed by MS-trained clinician was transcribed into NeurEx^TM^ App (Peter Kosa et al., 2018). NeurEx^TM^ computes traditional disability scales such as EDSS (Kurtzke, 1983), SNRS (Sipe et al., 1984) and others. We also extracted relevant subsystem scores of those neurological functions that, based on domain expertise, contribute to the spiral tracing (i.e., pyramidal and motor functions of hands, cerebellar functions, proprioception functions). Finally, we extracted semi-quantitative MRI data of CNS tissue destruction, focusing on brainstem, cerebellum, and medulla/upper cervical spinal cord. These are features previously validated as important in determining physical disability (Kelly et al., 2021; P. Kosa et al., 2015). The details of MRI sequences and computation of selected MRI features has been previously published (Kelly et al., 2021; P. Kosa et al., 2015).

Thus, together we tested 22 disability features in the MS training cohort (Table 2). Though spiral data were obtained from 22 HD, we note that clinical features were generated for only 9 HD (see Material and Methods for details).

### 2.4 Data processing and analysis

#### 2.4.1 Feature extractions

The raw sensor data was processed with signal and time series analysis methodologies to compute temporal, spatial, and spatiotemporal features. When appropriate, features were calculated following the work of (A. P. Creagh et al., 2020) in addition to some new features computed in this work and this generated a total of 40 spiral derived features (i.e., digital features). To measure temporal irregularities in the upper extremity function in neurological patients, previous research used speed and velocity as signals in the objective quantification of motor symptoms (Aghanavesi, Nyholm, Senek, Bergquist, & Memedi, 2017; Banaskiewicz, Rudzinska, Bukowczan, Izworski, & Szczudlik, 2009; A. P. Creagh et al., 2020; M. Memedi et al., 2015). Thus, we initially computed the velocity (v), radial and angular velocity (av) of the drawing spirals as follows:

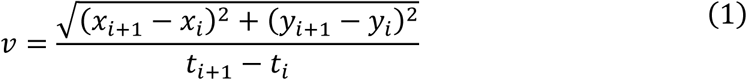

Where *x*_*i*_, *y*_*i*_, *i* = 1 … *N* are the horizontal and vertical coordinate of pixels on the screen respectively with *N* representing the total number of touch data points.*t*_*i*,_*i* = 1 … *N* is the timestamp converted to second. The radial velocity is computed as:

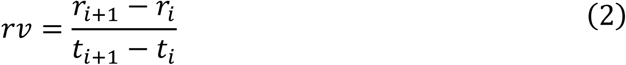

Where 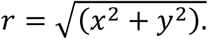. If we denote *θ* the four-quadrant inverse tangent 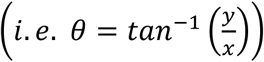, then the angular velocity takes the following form:

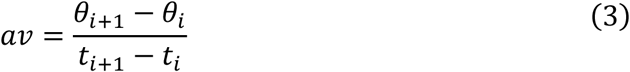

The sum, coefficient of variation, skewness, and kurtosis were computed for the velocity, radial velocity, and angular velocity respectively.

To calculate the degree of resemblance between the reference spiral and cohort’s drawing, we introduced features related to the Hausdorff distance, which quantify the extent to which each point in the reference spiral lies near the points in the cohort’s drawing following procedures illustrated in (A. P. Creagh et al., 2020; Dubuisson & Jain, 1994; Huttenlocher, Klanderman, & Rucklidge, 1993; Jeong & Srinivasan, 2017). Similar to Figure 3 in (Jeong & Srinivasan, 2017), a detail example procedure to calculate the Hausdorff distance of the reference and cohort’s drawing is presented in Figure 2.

**Figure 2.**
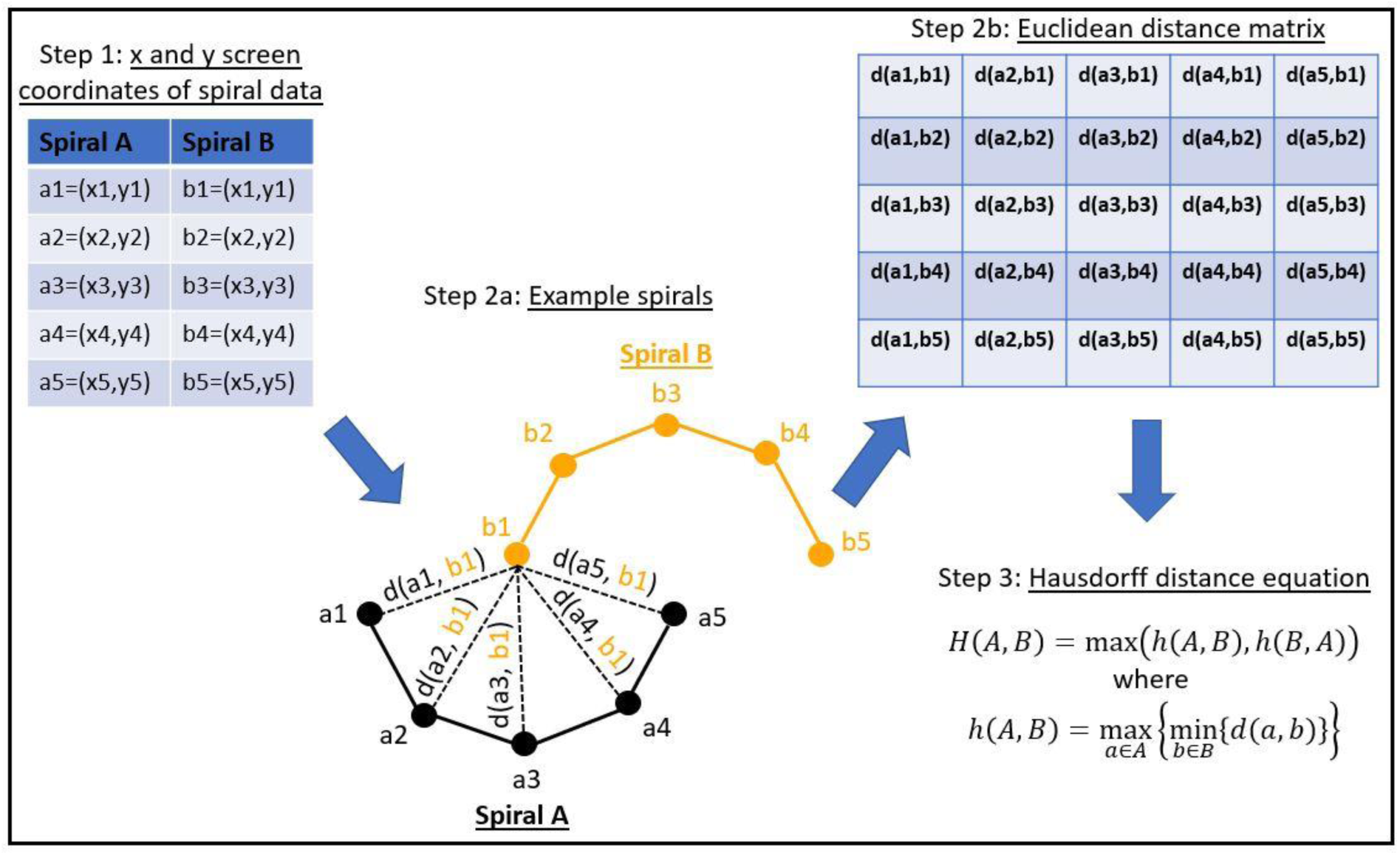
An example of the different steps to calculate the Hausdorff distance between two spiral curves. Step 1 represents an example of x and y coordinates reference spiral data (in orange; Spiral B) and cohort’s drawing data (in black; Spiral A). Step 2a is the Euclidean distance from b1 to each point in Spiral A denoted as d(ai, b1), i=1,2,3, 4, and 5. Dashed lines indicate the Euclidean distances. Step 2b represents the distance matrix from each point in Spiral B to each point in Spiral A. Step 3 finally provides the equation to calculate the Hausdorff distance after obtaining the distance matrix.

**Figure 3.**
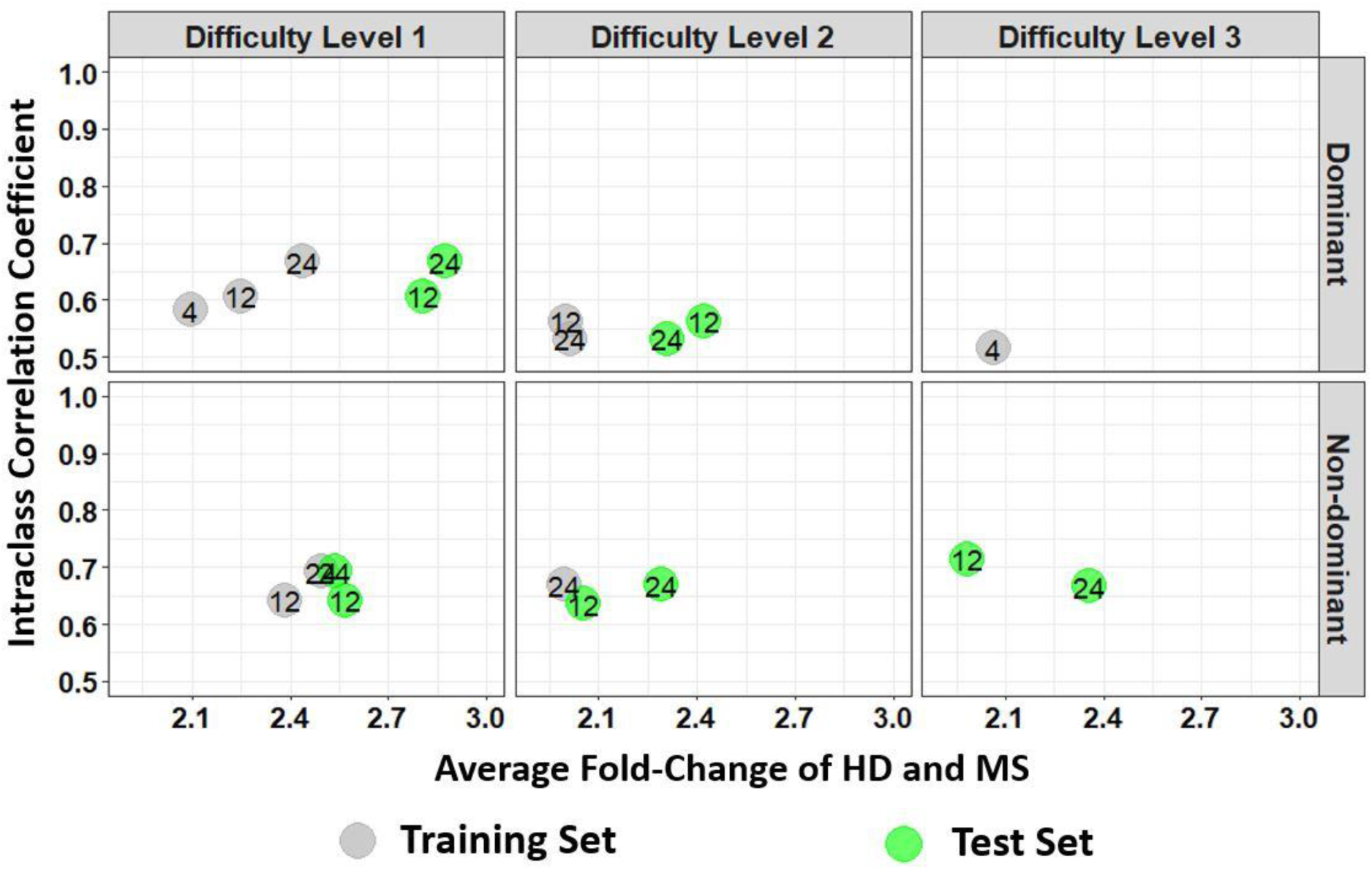
Average fold change of HD and MS of the spiral derived features with respect to their ICC where ICC was calculated from the granular data of the healthy donors (HD). The numbers indicate the feature’s labels as illustrated by the label in Table 2. Gray colors are features from the training set while green colors are the test set. Here we only include Fold-change > 2 and ICC > 0.5. A diagram of all features that are statistically significant between HD and MS in the training and test set for HD ICC is provided as Figures S5 and S6 respectively in the supplementary material. HD and MS denote Healthy Donors and Multiple Sclerosis respectively.

Prior to calculating the Hausdorff distance, the x and y screen coordinate points of the reference spiral were interpolated to the length of the cohort’s drawing’s coordinates using cubic spline interpolation (Fritsch & Carlson, 1980; Hyman, 1983; McKinley & Levine, 1998). Several Hausdorff distance related features were then calculated (e.g. maximum of Hausdorff distance, interquartile range of Hausdorff distance, etc.). All Hausdorff distance related features are provided in Table 3.

**Table 3.**
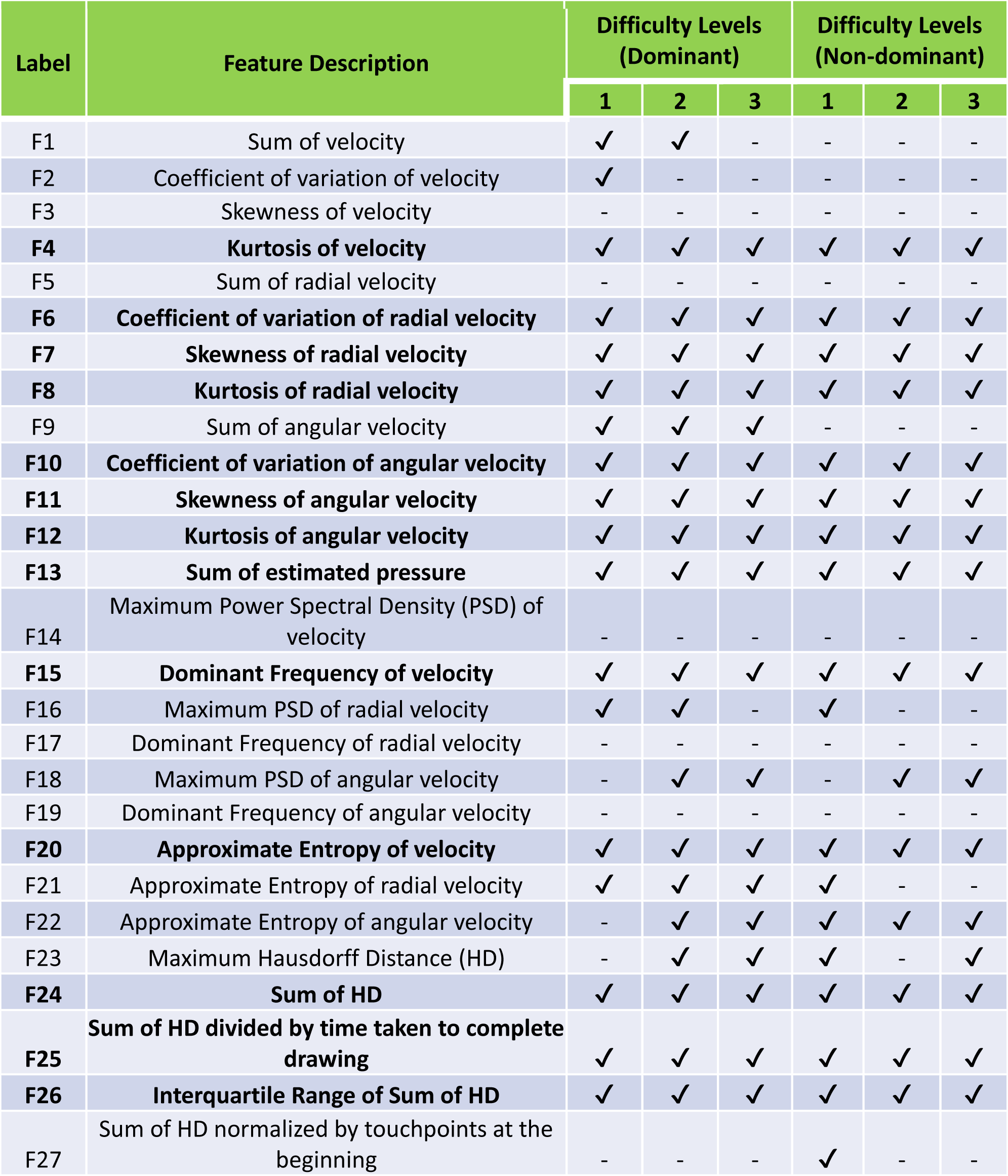
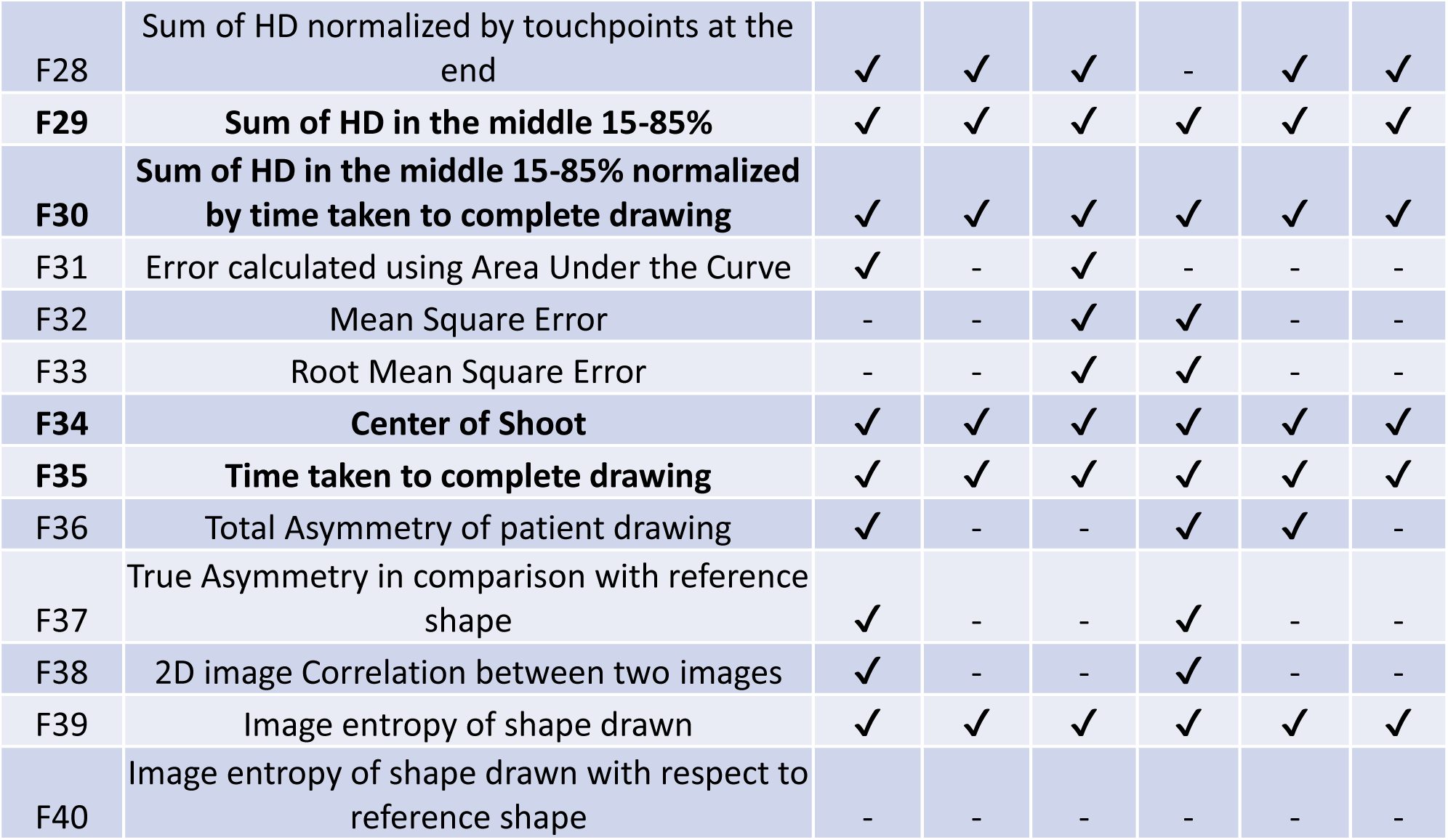
List of digital features relating to upper-extremity function calculated from the spiral drawing test. ✔ marks indicate features that are statistically significant different between HD and MS at the Benjamini-Hochberg (BH) adjusted p-value of 0.001. Features that are not statistically significant between HD and MS are marked with dashed symbols (-). The label and feature description of features that are statistically significant between HD and MS at both the dominant and non-dominant hands and at all difficulty levels are bolded. HD and MS denotes Healthy Donors and Multiple Sclerosis respectively.

| Label | Feature Description | Difficulty Levels (Dominant) |  |  | Difficulty Levels (Non-dominant) |  |  |
| --- | --- | --- | --- | --- | --- | --- | --- |
|  |  | 1 | 2 | 3 | 1 | 2 | 3 |
| F1 | Sum of velocity | ✓ | ✓ | - | - | - | - |
| F2 | Coefficient of variation of velocity | ✓ | - | - | - | - | - |
| F3 | Skewness of velocity | - | - | - | - | - | - |
| **F4** | **Kurtosis of velocity** | ✓ | ✓ | ✓ | ✓ | ✓ | ✓ |
| F5 | Sum of radial velocity | - | - | - | - | - | - |
| **F6** | **Coefficient of variation of radial velocity** | ✓ | ✓ | ✓ | ✓ | ✓ | ✓ |
| **F7** | **Skewness of radial velocity** | ✓ | ✓ | ✓ | ✓ | ✓ | ✓ |
| **F8** | **Kurtosis of radial velocity** | ✓ | ✓ | ✓ | ✓ | ✓ | ✓ |
| F9 | Sum of angular velocity | ✓ | ✓ | ✓ | - | - | - |
| **F10** | **Coefficient of variation of angular velocity** | ✓ | ✓ | ✓ | ✓ | ✓ | ✓ |
| **F11** | **Skewness of angular velocity** | ✓ | ✓ | ✓ | ✓ | ✓ | ✓ |
| **F12** | **Kurtosis of angular velocity** | ✓ | ✓ | ✓ | ✓ | ✓ | ✓ |
| **F13** | **Sum of estimated pressure** | ✓ | ✓ | ✓ | ✓ | ✓ | ✓ |
| F14 | Maximum Power Spectral Density (PSD) of velocity | - | - | - | - | - | - |
| **F15** | **Dominant Frequency of velocity** | ✓ | ✓ | ✓ | ✓ | ✓ | ✓ |
| F16 | Maximum PSD of radial velocity | ✓ | ✓ | - | ✓ | - | - |
| F17 | Dominant Frequency of radial velocity | - | - | - | - | - | - |
| F18 | Maximum PSD of angular velocity | - | ✓ | ✓ | - | ✓ | ✓ |
| F19 | Dominant Frequency of angular velocity | - | - | - | - | - | - |
| **F20** | **Approximate Entropy of velocity** | ✓ | ✓ | ✓ | ✓ | ✓ | ✓ |
| F21 | Approximate Entropy of radial velocity | ✓ | ✓ | ✓ | ✓ | - | - |
| F22 | Approximate Entropy of angular velocity | - | ✓ | ✓ | ✓ | ✓ | ✓ |
| F23 | Maximum Hausdorff Distance (HD) | - | ✓ | ✓ | ✓ | - | ✓ |
| **F24** | **Sum of HD** | ✓ | ✓ | ✓ | ✓ | ✓ | ✓ |
| **F25** | **Sum of HD divided by time taken to complete drawing** | ✓ | ✓ | ✓ | ✓ | ✓ | ✓ |
| **F26** | **Interquartile Range of Sum of HD** | ✓ | ✓ | ✓ | ✓ | ✓ | ✓ |
| F27 | Sum of HD normalized by touchpoints at the beginning | - | - | - | ✓ | - | - |
| F28 | Sum of HD normalized by touchpoints at the end | ✓ | ✓ | ✓ | - | ✓ | ✓ |
| F29 | **Sum of HD in the middle 15-85%** | ✓ | ✓ | ✓ | ✓ | ✓ | ✓ |
| F30 | **Sum of HD in the middle 15-85% normalized by time taken to complete drawing** | ✓ | ✓ | ✓ | ✓ | ✓ | ✓ |
| F31 | Error calculated using Area Under the Curve | ✓ | - | ✓ | - | - | - |
| F32 | Mean Square Error | - | - | ✓ | ✓ | - | - |
| F33 | Root Mean Square Error | - | - | ✓ | ✓ | - | - |
| F34 | **Center of Shoot** | ✓ | ✓ | ✓ | ✓ | ✓ | ✓ |
| F35 | **Time taken to complete drawing** | ✓ | ✓ | ✓ | ✓ | ✓ | ✓ |
| F36 | Total Asymmetry of patient drawing | ✓ | - | - | ✓ | ✓ | - |
| F37 | True Asymmetry in comparison with reference shape | ✓ | - | - | ✓ | - | - |
| F38 | 2D image Correlation between two images | ✓ | - | - | ✓ | - | - |
| F39 | Image entropy of shape drawn | ✓ | ✓ | ✓ | ✓ | ✓ | ✓ |
| F40 | Image entropy of shape drawn with respect to reference shape | - | - | - | - | - | - |

Two approaches were utilized to compute the error related features between the reference spiral and the cohort’s drawings. The first error was computed using the trapezoid to integrate over the two spiral regions. This error was calculated by finding the intersection of the two-spiral region (i.e., the difference between the two areas in magnitude). For instance, we note the following trapezoidal formula from (Aghanavesi et al., 2017):

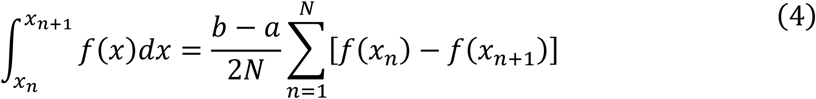

Where *N* is the total number of x or y screen coordinate points and 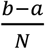 is the spacing between points. Suppose the reference spiral and the cohort’s spiral are denoted by *f*_*ref*_(*x*, *y*) and *f*_*co*ℎ_(*x*, *y*) respectively. Then the error based on the trapezoidal rule becomes

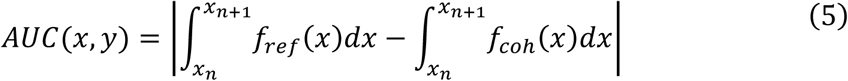

Where |. | is the absolute value of the difference between the two Area Under the Curve (AUC). We now proceed with the second error calculated using the following 2D Mean Square Error (MSE; (Asamoah, Ofori, Opoku, & Danso, 2018)):

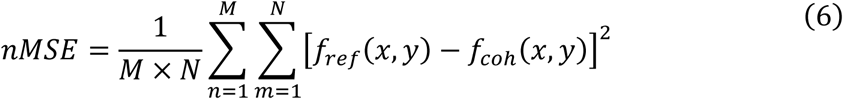

Where *M* and *N=2* are the number of data points and coordinates points respectively. Again, a spline interpolation was used on the cohort’s data point to the M length of the reference data point prior to error calculation. Furthermore, to obtain the similarity between the reference spiral and cohort’s drawing, the following 2D correlation coefficient from (Aljanabi, Hussain, & Lu, 2018) was utilized:

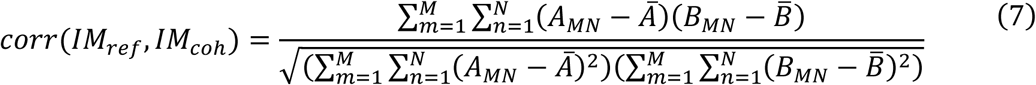

Where *A_MN_* and *B_MN_* are the reference and cohort’s spiral coordinate points with dimension *M x 2* respectively. 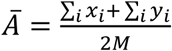 and 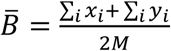 are the reference and cohort’s spiral mean. Note that the two spirals mean are not necessarily equal as the x and y coordinate points differ. A full list of features calculated are provided in Table 3. When applicable, references of the calculated spiral derived features are provided in Table S1 in the supplemental material.

#### 2.4.2 Statistical analysis

Statistical analyses were used to evaluate the validity and strength of features (i.e., clinical disability scales and spiral derived features) on assessing the upper extremity function in MS patients. The analyses were conducted using the R software (R Version 4.0.4; (Team, 2021)). Recall that the MS datasets were separated into a 2/3 training and 1/3 test set weighted by the average 9HPT disability scale. A cutoff Benjamini-Hochberg (BH; (Benjamini & Hochberg, 1995)) adjusted p-value < 0.001 was used to establish statistically significant differences for comparing the HD and MS cohorts. All features that were not statistically significant using the unpair Two-Samples Wilcoxon test (Wilcoxon, 1945) in the training set were removed from subsequent analysis. Moreover, average fold change (FC) between the HD and MS was computed at all difficulty levels and at both dominant and non-dominant hand. A FC > 2 was used as cutoff of significant difference between HD and MS.

Test-Retest reliability of the spiral derived features were measured using the Intraclass Correlation Coefficient (Andrea Vianello, Luca Chittaro, Stefano Burigat, & Riccardo Budai) of features obtained from the granular testing HD and MS sub-cohorts. As stated by (Koo & Li, 2017; WEIR, 2005), there are several versions of the ICC that can give different results when used on the same dataset. However, the authors pointed that the two-way mixed-effects model and the absolute agreement are more appropriate for test-retest reliability studies. Thus, an ICC with two-way mixed-effects model and the absolute agreement was used in this study. Following the recommendation of (Koo & Li, 2017) that stated that an ICC between 0.5 and 0.75 are consider moderate, a spearman correlation matrix between clinical disability scales (see Table 2) and spiral derived features (see Table 3) with ICC > 0.5 and FC > 2 were constructed. A BH adjusted p-value > 0.05 was used to access statistical significance of the correlation test.

To determine the existing relationship between the significant spiral derived features (i.e. HB adjusted p-value < 0.001, FC > 2, and ICC > 0.5 between HC and MS) and the statistically significant clinical disability scales, four different regression models (Elastic Net or ElasticNet, Support Vector Regression with Radial Basis Function Kernel or SVR Radial, Random Forest or RF, and Stochastic Gradient Boosting or GBM) were used where the clinical disability scale were the dependent variables while spiral derived features were the independent variables. For all regression models, the ‘caret’ library (Kuhn, 2008) in the R software was used along with other libraries such as ‘glmnet’ (Friedman, Hastie, & Tibshirani, 2010) for ElasticNet model, ‘randomForest’ (Liaw & Wiener, 2002) for RF model, and ‘xgboost’ (Chen et al., 2019) for GBM model. Prior to the regression modeling, outliers in the spiral derived features were identified as feature values that lie outside of ± 2(q_0.9_ – q_0.1_) where q_p_ is the p-quantile (Hyndman et al., 2020). To reduce variability in the features, all variables were bi-symmetric log transformed using the transformation formula *y* = *sgn*(*x*)*log*_10_(1 + |*x*/*C*|) where *y* is the transformed function of the *x* variable, C has a default value of 1/ln(10), and *sgn(x)* is the mathematical Signum function (as presented in (Webber, 2012)).

Moreover, a linear regression model was used to assess the relationship between selected clinical disability scales and the sum of the Hausdorff distance (Feature F24 in Table 3). During the analysis, adherence to the normality assumptions of the residuals was tested using histograms and quantile plots. All models were evaluated using the Root Mean Square Error (RMSE; measured in seconds) and the coefficient of determination (R-square) of the prediction. Apart from the linear regression model, all model parameters (i.e. the penalty strength parameter λ and the penalties from both L1 and L2 regularization parameter α in ElasticNet; the cost value C and γ of the SVR Radial; the number of variables randomly sampled at each split in the RF model; the number of tree, the interaction depth, the minimum number of samples in tree terminal nodes, and the learning rate in GBM model) were tuned via grid search. 5-fold cross-validation (CV) with 10 repetitions was used to assess the model suitability in the training cohort. The out-of-sample test performance was evaluated using the final model from the 5-fold CV based on the RMSE to predict clinical disability scales given the test datasets (i.e., independent validation cohort).

## 3. Results

### 3.1 Features evaluation

To determine clinical disability scales and spiral derived features that are relevant for further analysis, statistical significance between HD and MS was calculated using unpair Two-Samples Wilcoxon test. With the exception of four features (NeurEx^TM^ vision score, brainstem atrophy, medulla/upper c-spine atrophy, and cerebellum atrophy), all clinical disability scales and MRI features were found to be statistically significant at p < 0.001 after adjusting for the p-value using BH approach (Table 2). In the dominant hand category, 28, 26, and 27 spiral derived features were statically significant (BH adjusted p-value < 0.001) at the difficulty level 1, 2, and 3, respectively. However, in the non-dominant hand category, 28, 22, and 22 spiral derived features were statically significant at the difficulty level 1, 2, and 3, respectively. While spiral derived features that were statistically significant between HD and MS varies between dominant, non-dominant hands and difficulty levels, 19 of these features were consistently statistically significant at all levels and both hands (see bold feature description in Table 3).

Furthermore, a look at the FC of HD and MS with respect to the ICC indicated that only 3 spiral derived features (kurtosis of velocity, kurtosis of angular velocity, and sum of the Hausdorff Distances) have FC > 2 and ICC > 0.5 when ICC was calculated using the granular data of the healthy donors (Figure 3). When ICC was computed for the MS patients, kurtosis of radial velocity, kurtosis of angular velocity, and sum of Hausdorff Distance have FC > 2 and ICC > 0.5 (Figure 4). In general, four spiral derived features (i.e., kurtosis of velocity, kurtosis of radial velocity, kurtosis of angular velocity, and sum of Hausdorff Distance) were found to be statistically significant between HD and MS, have FC > 2, and have healthy donors or MS patients ICC > 0.5. These features have a moderate strength of test-retest reliability and are significantly different in HD and MS as indicated by their FC and ICC (Figure 3 and Figure 4). The difference of HD and MS patients in selected clinical features and the four most impactful spiral derived features was depicted by Boxplots (See Figure S1 for Boxplot of selected clinical features, Figures S2-S4 for Boxplots of most impactful spiral derived features at the difficulty level 1, 2, and 3 respectively in the supplemental material). Most spiral-tracing features have median value higher in the MS as compared to the HD (Figures S1-S4). Generally, ICC of MS patients are also higher than ICC of HD (Figure 5). This is expected based on higher inter-individual variance of spiral-tracing outcomes in MS vs HD.

**Figure 4.**
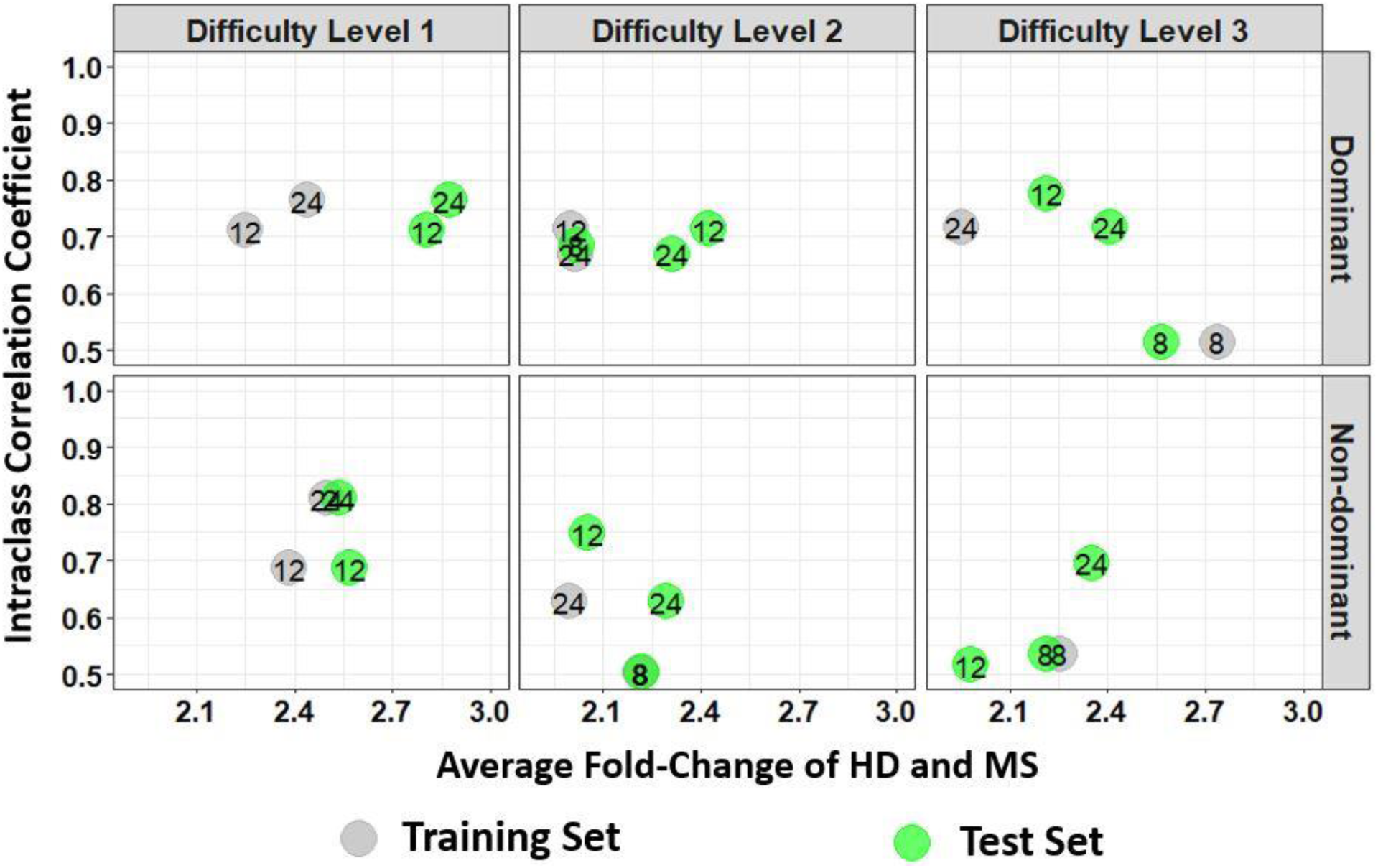
Average fold change of HD and MS of the spiral derived features with respect to their ICC where ICC was calculated from the granular data of the MS patients. The numbers indicate the feature’s labels as illustrated by the label in Table 2. Gray colors are features from the training set while green colors are the test set. Here we only include Fold-change > 2 and ICC > 0.5. A diagram of all features that are statistically significant between HD and MS in the training and test set for MS patients ICC is provided as Figures S7 and S8 respectively in the supplementary material. HD and MS denote Healthy Donors and Multiple Sclerosis respectively.

**Figure 5.**
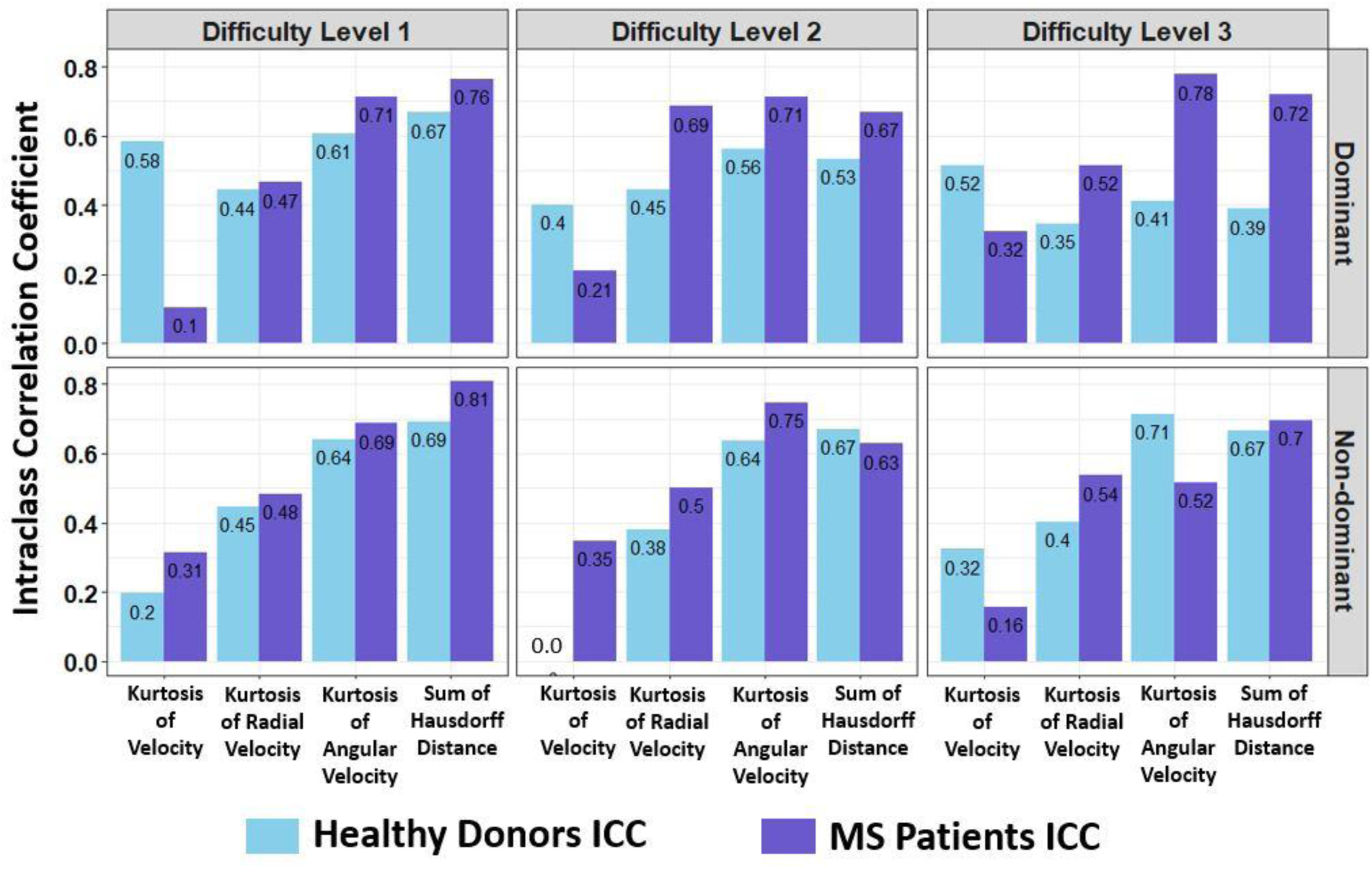
Comparison of the Intraclass Correlation Coefficient calculated from the granular data of the healthy vs. MS cohorts. The x-axis represents spiral derived feature with Fold-change > 2 and ICC > 0.5. MS denotes Multiple Sclerosis.

To determine the relationship between the clinical disability scales and most impactful spiral derived features (Kurtosis of velocity, radial velocity, angular velocity, and the sum of Hausdorff distances), a spearman rho correlation matrix among the features was constructed (see Figure 6 for difficulty level 1 and 2, and Figure S9 for difficulty level 3).

**Figure 6.**
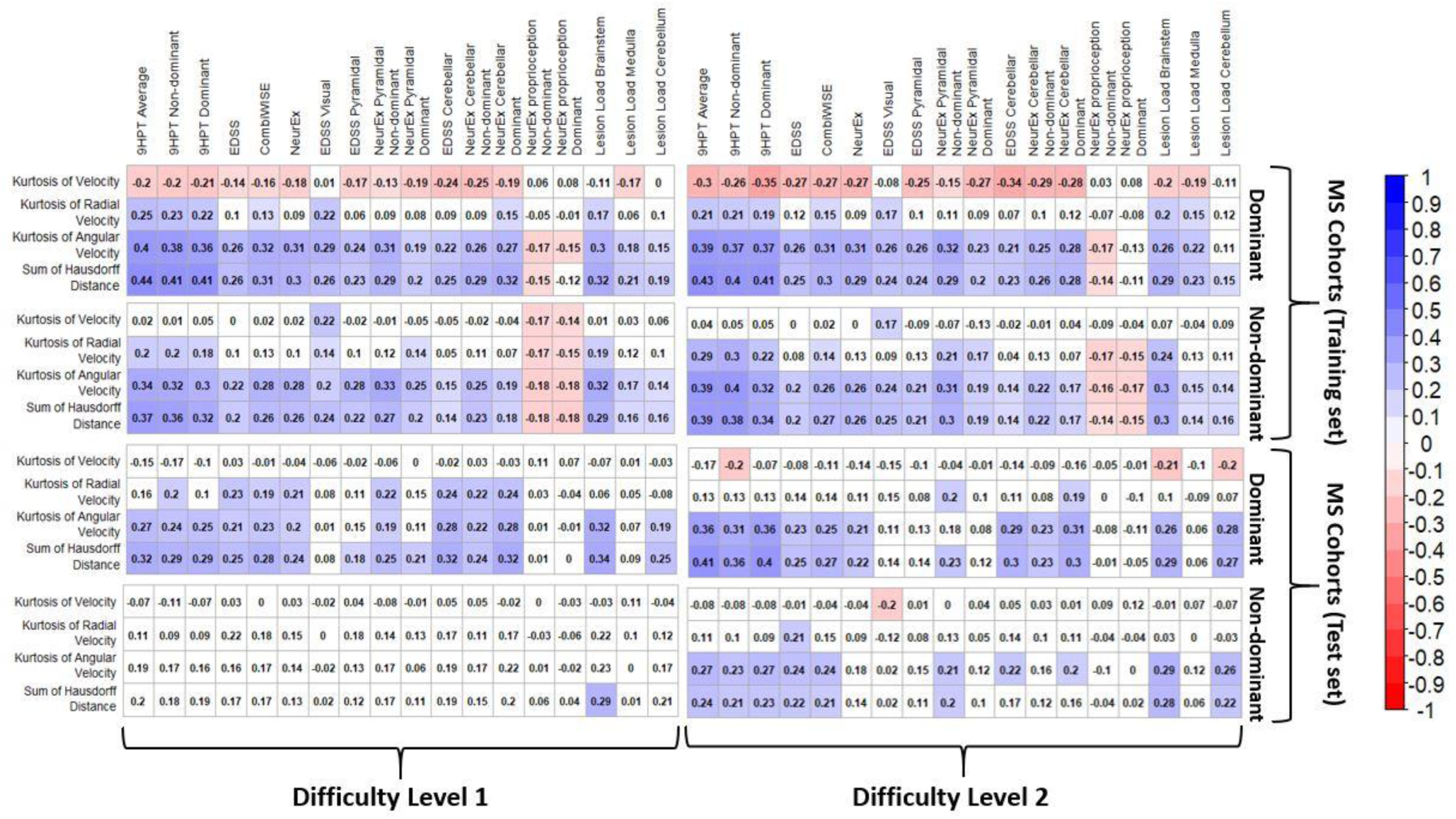
Spearman rho correlation matrix between the statistically significant clinical disability and the top four most significant spiral derived features based on FC at the difficulty level 1 and 2. The number indicates the spearman correlation coefficient. Red is negative correlation while blue stand for positive correlation. The white color indicates correlation that are not statistically significant at BH adjusted p-value of 0.05.

In general, the highest correlations were seen among the 9HPT Average, EDSS, CombiWISE, NeurEx^TM^ and the spiral derived features at the dominant hand levels (Figure 6 and Figure S9). Among the spiral derived features, the sum of the Hausdorff distances had the highest correlations in both cohorts, but the strength of correlations was weak to moderate.

From the neurological examination subdomains only motoric and cerebellar functions (but not proprioception) reliably correlated with best spiral tracing features.

We observed a very high positive correlation between the sum of Hausdorff distances and the time taken to complete the drawing in both the HD and MS patients (Figures S10-S12; spearman rho > 0.97 for both dominant and non-dominant hand and at all difficulty levels). This is counter intuitive as we expected that increasing speed of spiral drawing will negatively affect tracing accuracy. Instead, it appears that disability and/or person’s confidence of his/her ability to trace the spiral affected both speed and accuracy of the tracing congruently.

### 3.2 ML models of best spiral tracing features and their independent cohort validation

Four regression models (ElasticNet, SVR Radial, RF, GBM) were used to evaluate the relationship between clinical disability scales and our four most impactful features (Kurtosis of velocity, Kurtosis of Angular velocity, and Sum of the Hausdorff Distances). The models had the best performance predicting the clinical disability scales in the (small) HD cohort based on the mean RMSE and R-square across the 5-Fold CV with 10 repetitions (see Tables S2, S5, and S8 for difficulty level 1, 2 and 3 respectively in the supplemental material). Among the HD at the difficulty level 1, RF model performed the best by explaining at most 81 percent in clinical disability scales when the dependent variables were 9HPT Average or EDSS (Table S2). When the dependent variables are CombiWISE or NeurEx, GBM had the best performance at the difficulty level 1 with R^2^ value between 0.72 and 0.90 (Table S2). The results of the percent variance explained in model outcomes at the difficulty levels 2 and 3 in healthy donors were lower compare the difficulty level 1 but still range between 30 and 75 percent (Table S5 and S8 in the supplemental material).

Model’s performance in the (much larger) MS training cohort were much lower at all difficulty levels in comparison to results from the HD. The SVR Radial performed the best by explaining only 6 to 23 percent of variance in clinical disability scale depending on the hand used (dominant or non-dominant), difficulty levels, and clinical disability scale (see Tables S3, S6, and S9 for difficulty level 1, 2 and 3 respectively in the supplemental material).

However, the out-of-sample test performance (i.e., the independent validation cohort) were much lower compare to the result from the 5-fold CV of the training cohort (see R^2^ values between 0.0002 and 0.18 in Tables S4, S7, and S10 for difficulty level 1, 2 and 3 respectively in the supplemental material). Of these, models with 9HPT Average yield the best performance with an R-square between 0.10 and 0.18 but only for dominant hand (Tables S4, S7, and S10 for difficulty level 1, 2 and 3 respectively in the supplemental material). However, in the independent validation cohort the RSV models validated the worst.

Thus, we conclude that cross-validation of the training cohort is overly optimistic and does not reliably predict an independent validation cohort performance.

Given that the sum of Hausdorff distances had the highest correlation with the clinical disability scale at all difficulty levels (Figure 6 and Figure S9), a linear regression models were constructed to measure the relationship between the disability scales and the sum of Hausdorff distances alone. In general, all clinical disability scales were positively correlated with the sum of Hausdorff Distances (see Figure 7 for correlation with Average 9HPT and S13-S15 for correlations with EDSS, CombiWISE, and NeurEx respectively). The validation cohort performance of the sum of Hausdorff Distances alone (Figure 8 for 9HPT) was comparable to the more complex ML-based models (i.e., R^2^ between 0.04 and 0.14 in Table S11).

**Figure 7.**
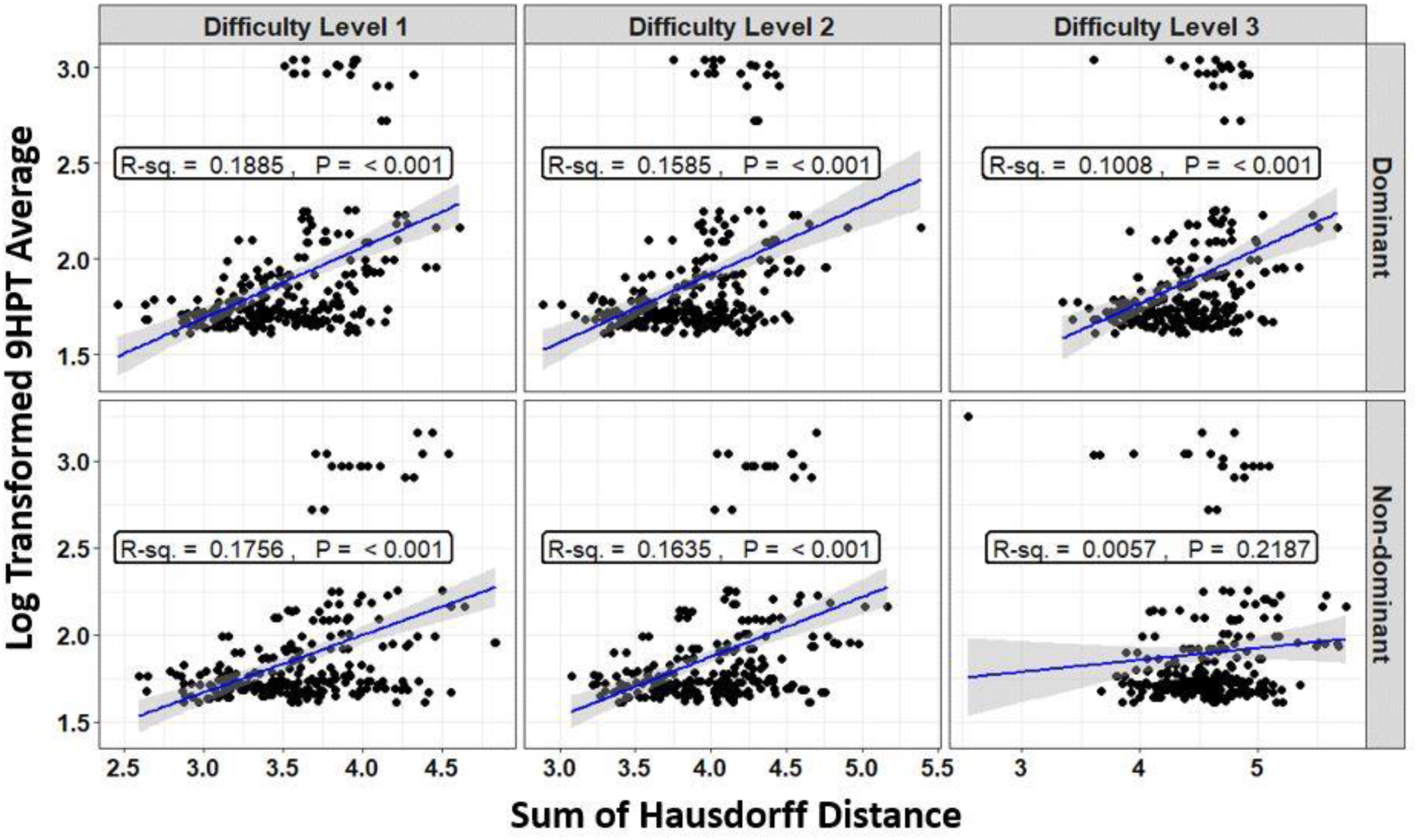
Relationship between the sum of the Hausdorff distances and the 9HPT Average (in seconds) of the MS cohorts in black dots. Regression lines are shown in solid blue line while the gray shaded area constitute the 95% confidence interval associated with the mean model’s prediction. The R-square denoted R-sq. indicates the percent of variance in the 9HPT Average that can be explained by the sum of Hausdorff distances. P is the model’s p-value. MS denotes Multiple Sclerosis.

**Figure 8.**
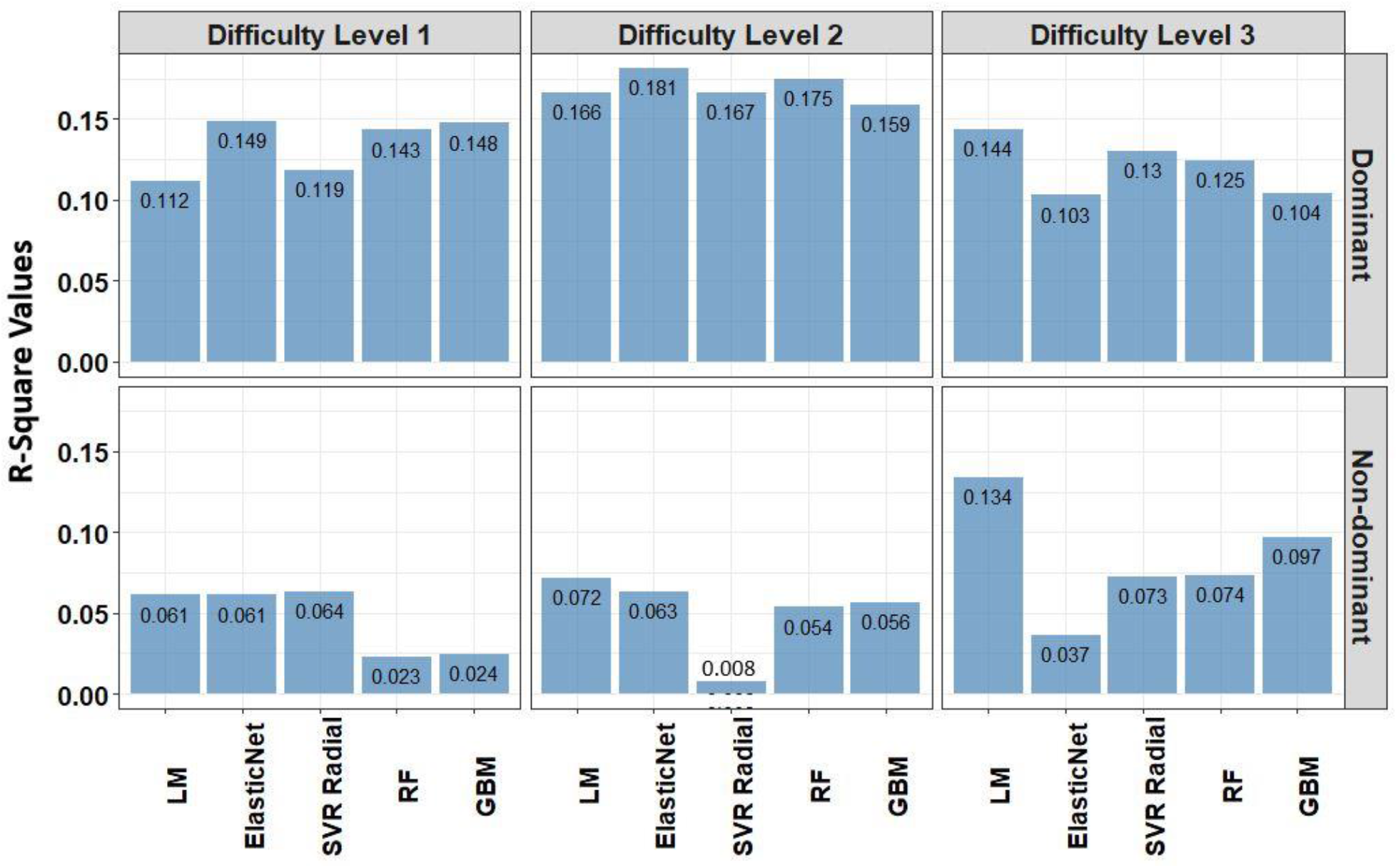
The out-of-sample test performance (i.e., independent validation) of the predictive regression models when 9HPT Average (in seconds) is the response variable and the sum of Hausdorff Distances is the only explanatory variable in the linear regression model (LM). The explanatory variables in the Elastic Net (ElasticNet), Support Vector Regression with Radial Basis Function kernel (SVR Radial), Random Forest (RF), and Stochastic Gradient Boosting (GBM) models are Kurtosis of Velocity, Kurtosis of Radial Velocity, Kurtosis of Angular Velocity, and the sum of Hausdorff Distances. The test performance was measured using R-square (R^2^) of model predictions per dominant and non-dominant hands among the MS patients at the difficulty level 1, 2, and 3. MS denotes Multiple Sclerosis.

Overall, we observed better predictive accuracy (based on R-square) in dominant hand category than non-dominant hand category and for difficulty levels 1 and 2 compared to difficulty level 3 in the dominant hand (Figure 8).

## 4. Discussion

Test reproducibility, (measured as ICC, signal-to-noise ratio or concordance coefficient) is generally given lesser importance in the test design than test sensitivity. Presented analyses of the Spiral tracing support the notion that test reproducibility is essential determinant of its clinical utility. Achieving high reproducibility of digital tests should be at forefront of the medical app developers.

Spiral tracing is complex test that includes many neurological functions: fine finger movements, negatively affected by motoric dysfunction, proprioceptive loss and cerebellar dysfunction, eye-hand coordination, affected by vision, oculomotor and cerebellar dysfunctions and cognition or anxiety associated with anticipated test difficulty. Additionally, the precision of the test is affected by time of test execution, even though we observed, counterintuitively, strong *positive* correlation between measures of test accuracy such as sum of Hausdorff distance and the time it took to perform the tracing, even in HD. This suggests that time was not the primary driver of the inaccuracy of tracing. Rather, combined neurological disability and/or lack of confidence in the ability to perform test both fast and accurately, negatively affected the test performance. This explains why Spiral tracing features that adjusted accuracy of tracing for the velocity performed worse than the most successful accuracy measure, the sum of Hausdorff distance.

The comparison of cross-validation performance of the MS training cohort with the performance of the ML-based models in the true independent validation cohort demonstrated that training cohort cross-validation performance over-estimates performance of the test in subjects who did not contribute to the model development: when the performance of the strongest ML-based models (i.e., modeling 9HPT) are compared between cross-validation of the training cohort and the independent validation cohort, all four ML algorithms greatly over-estimated performance of the models in the independent validation cohort. In fact, the best feature of the Spiral tracing (the sum of Hausdorff distances) performed comparatively to the ML-based models in the independent validation (Figure 8). This overestimation of the model performance from the training cohort data, even when training cohort results are based on cross-validation, is the rule we observed uniformly in the past decade of our experience with independent validation of complex models. In fact, we used to not even show the training cohort results in our publications, as we consider them irrelevant. However, after realizing that vast majority of ML studies in biomedical literature do not use independent validation cohort and that most readers and reviewers consider cross-validation of the training cohort equivalent to the independent validation, we now routinely publish training cohort data to demonstrate the level of overfit in comparison to truly independent validation cohort.

We also point out that the cross-validation performance of the training cohort does not faithfully predict even the ranking of the models: e.g., the RSV models explained the highest proportion of variance in the training cohort cross-validation, but performed especially poorly in the independent validation cohort.

In this regard, because COVID19 pandemic precluded us from recruiting independent validation cohort of HD, we consider cross-validation performance of the HD models unrealistically optimistic (especially in view of the small number of HD) and fully expect that those models represent overfit. Therefore, the HD models should not be considered promising without an independent validation.

The poor performance of these ML-based models was, in our experience, expected based on poor ICCs and weak univariate correlations of individual Spiral tracing features with the gold standard of neurological disability measures and brain MRI markers of CNS injury in MS cohorts. In comparison, much simpler test such as rapidly tapping on the screen of the smartphone correlated much stronger with analogous disability measures (i.e., up to spearman Rho of 0.76) and were also much more intra-individually stable(Alexandra K. Boukhvalova et al., 2018). Interestingly, both 9HPT and smartphone finger tapping differentiated MS from HD better for non-dominant hand, observation that was reproduced for 9HPT in multiple studies (A. K. Boukhvalova et al., 2018; P. Kosa et al., 2016; Tanigawa et al., 2017). We interpreted this observation by functional repair: even though MS likely affects both hands equally, the daily use of dominant hand promotes repair, both as remyelination and establishment of new synaptic circuits by remaining neurons. Therefore, digital tests of non-dominant hand, which has less rehabilitation/repair, are more sensitive to measure difference between MS patients and HD and to measure progression of disability in time. Surprisingly, non-dominant performed much worse in Spiral tracing test, in both MS cohorts. We believe that this was due to higher intra-individual variance/greater noise, that has little to do with disability and more to do with test complexity. In fact, as test complexity increased to Level 3, the reproducibility and clinical relevance of the Spiral tracing features decreased quite dramatically.

We recognize that poor reproducibility of the Spiral tracing observed in our study may be mitigated in situations where spiral tracing is performed on tablets and therefore, the spiral is much larger (Erasmus et al., 2001). We developed NeuFun-TS for smartphones rather than tablets, due to larger World-wide prevalence and greater availability of different sensors in the former compared to the latter. Clearly, the test selection must consider the screen size difference for apps targeting different mobile devices.

In conclusion, in self-administered digital measurements of neurological functions, the designers should strive to develop tests that are easy to perform and therefore highly reproducible, but still reflect a specific neurological (dys)function. These simpler tests will likely be (by design) less sensitive than tests that depend on multiple neurological functions, but the sensitivity can be restored by aggregating results from multiple simple tests, as is being done in NeuFun-TS. However, the total time necessary to complete all tests in NeuFun-TS will likely determine compliance with longitudinal testing. Therefore, as Spiral tracing does not add clinical value beyond existing tapping (Tanigawa et al., 2017), balloon popping (Alexandra K. Boukhvalova et al., 2018) and level tests (Boukhvalova et al., 2019), we plan to drop Spiral tracing from NeuFun-TS standard tests. Spiral tracing Fourier analysis to identify tremor, its frequency and severity may still be very useful in patients with movement disorders.

## Data Availability

The datasets analyzed in this study along with the R Code have been made available at https://github.com/bielekovaLab/Bielekova-Lab-Code/tree/master/FormerLabMembers/Messan_Komi

## 6. Author Contributions

BB conceived the study design. BB, LP, TH, YK, VM, and PK performed the app construction and clinical data collection. BB and KSM conceived the analytical methodologies. KSM performed the statistical analysis and ML modelling with their respective visualizations. All authors participated in writing and editing the manuscript.

## Acknowledgements

We would like to thank clinicians Alison Wichman and Mary Sandford, research nurse Tiffany Hauser, and patient care coordinator Michelle Woodland for their excellent patient care. We would like to thank former laboratory members Alex Boukhvalova, Ann Weideman, Joshua Milstein, Jonathan Phillips, Kayla Jackson, Erin Kelly, Paavali Hannikainen, and Samuel Wachamo for helping with data collection in the clinic. Finally, we would like to thank our patients, their caregivers, and healthy volunteers for being partners in research – without them this work would be possible.

## Supplemental Material

**Figure S1.**
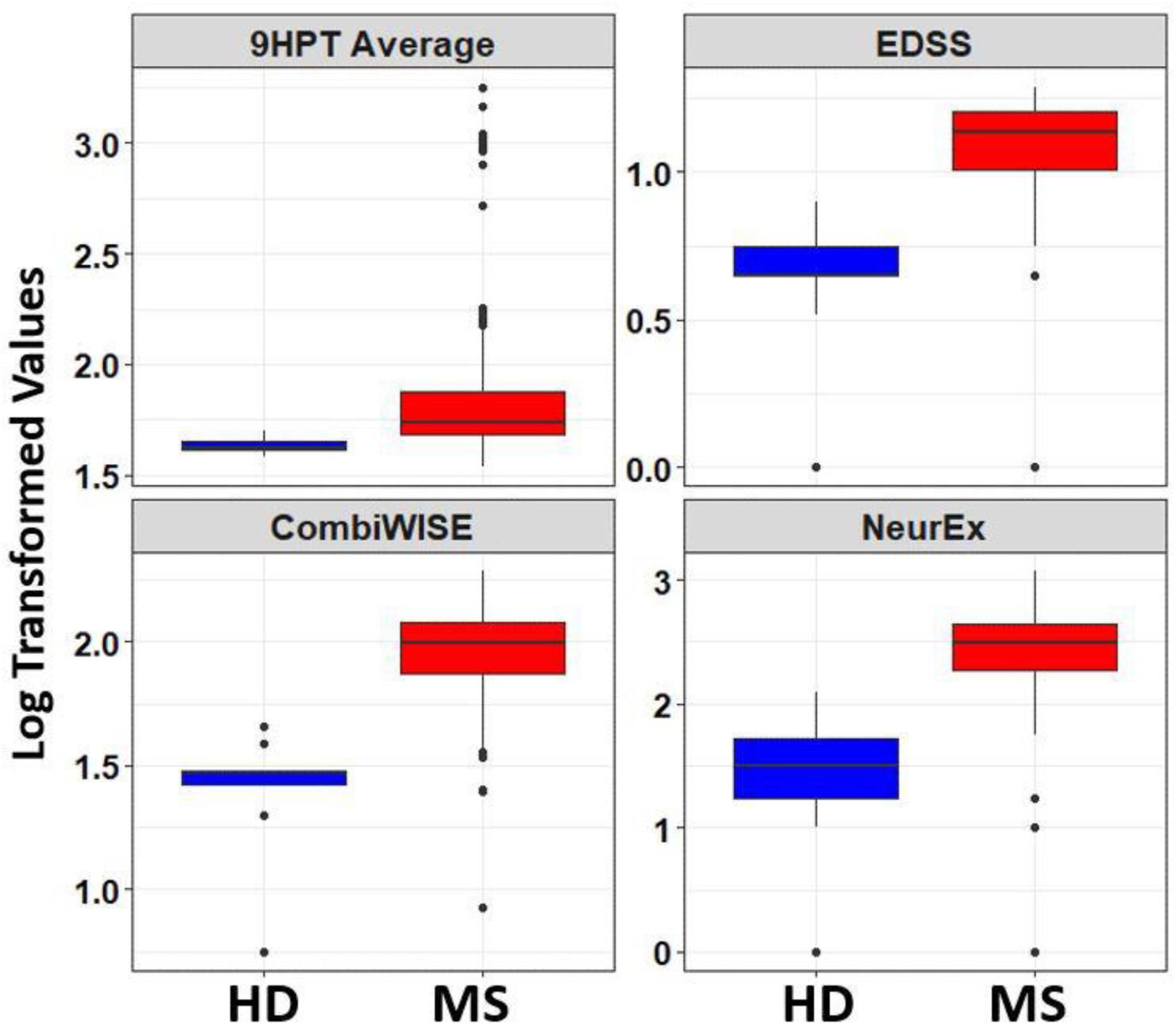
Boxplot of selected clinical disability scales with respect to healthy donors (HD) and MS patients. The blue color indicates the healthy donors while red shows the MS patients. Values are shown using bi-symmetric log transformation. MS denotes Multiple Sclerosis.

**Figure S2.**
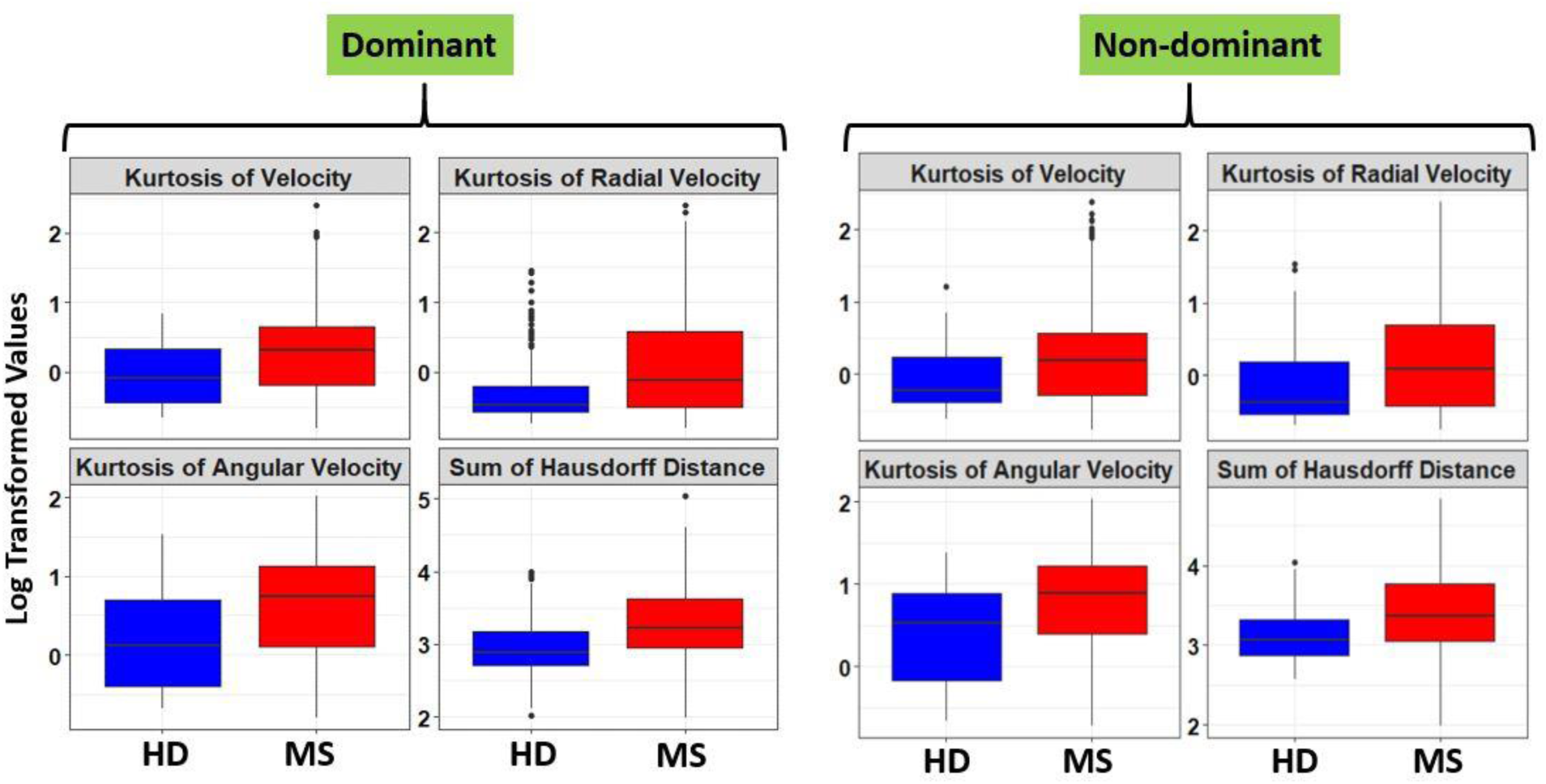
Boxplot of selected spiral derived features with Fold-Change > 2 and ICC > 0.5 with respect to healthy donors (HD) and MS patients at the difficulty level 1. The blue color indicates the healthy donors while red shows the MS patients. Values are shown using bi-symmetric log transformation for both the dominant and non-dominant hand. MS denotes Multiple Sclerosis.

**Figure S3.**
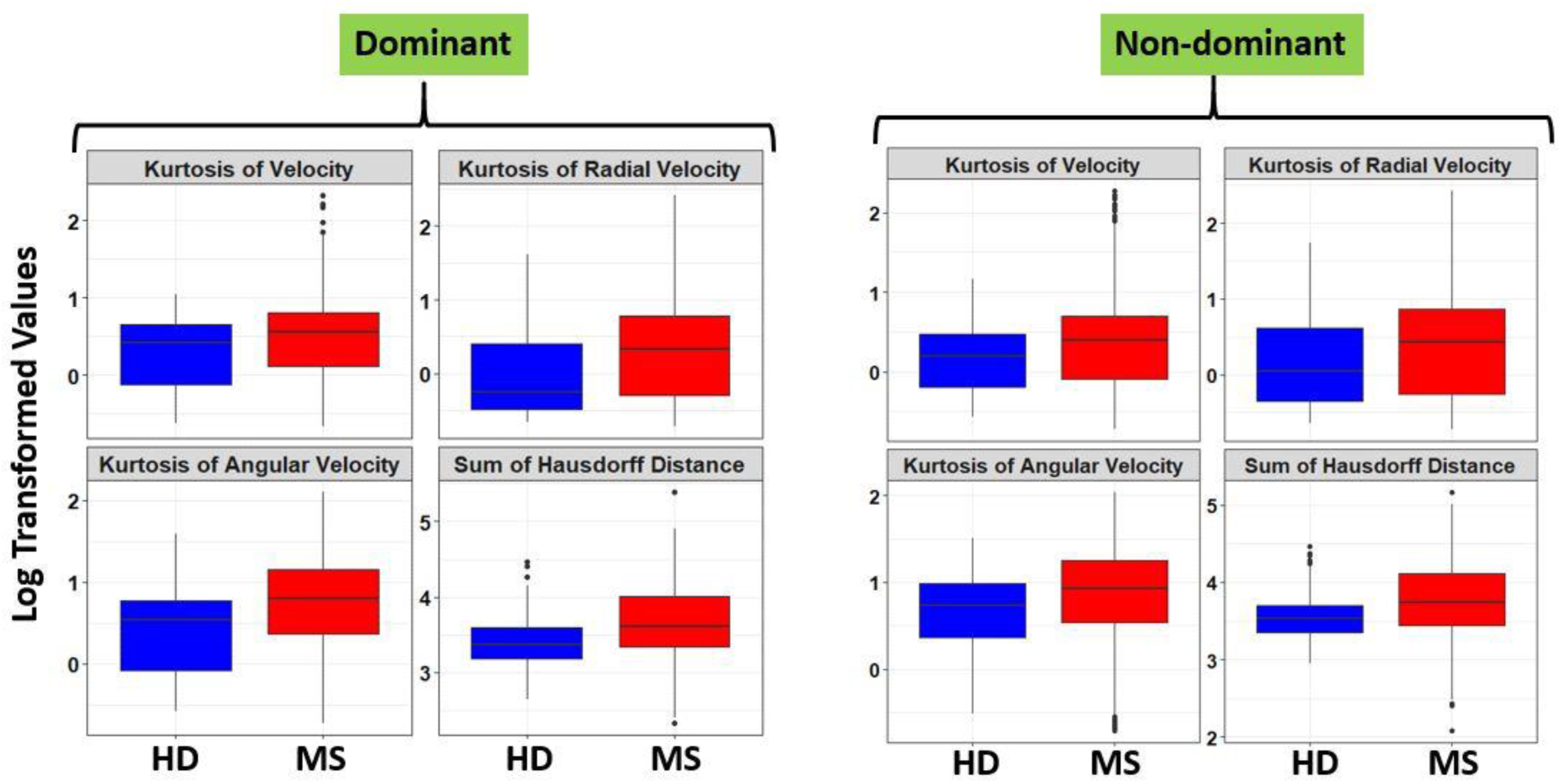
Boxplot of selected spiral derived features with Fold-Change > 2 and ICC > 0.5 with respect to healthy donors (HD) and MS patients at the difficulty level 2. The blue color indicates the healthy donors while red shows the MS patients. Values are shown using bi-symmetric log transformation for both the dominant and non-dominant hand. MS denotes Multiple Sclerosis.

**Figure S4.**
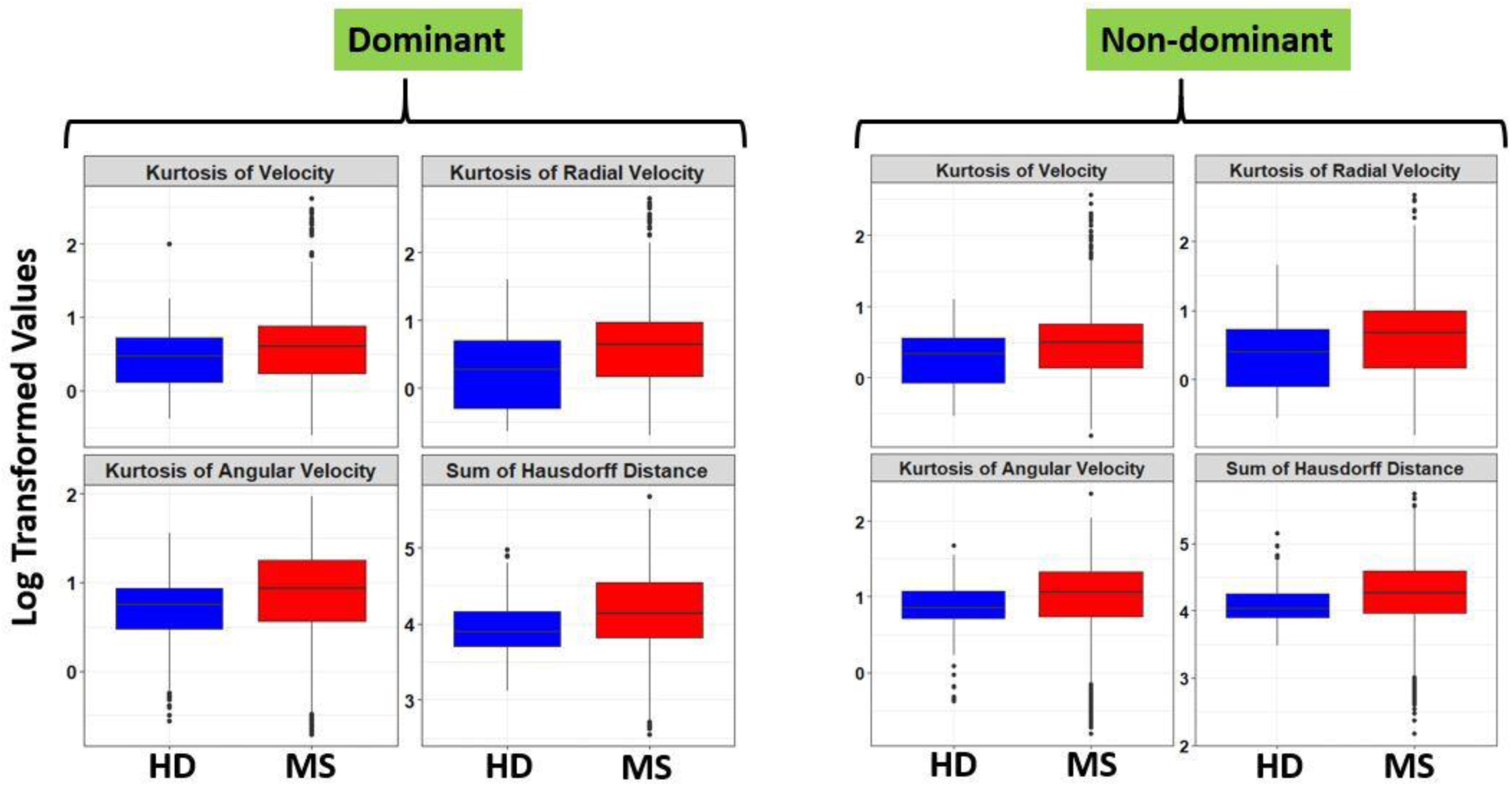
Boxplot of selected spiral derived features with Fold-Change > 2 and ICC > 0.5 with respect to healthy donors (HD) and MS patients at the difficulty level 3. The blue color indicates the healthy donors while red shows the MS patients. Values are shown using bi-symmetric log transformation for both the dominant and non-dominant hand. MS denotes Multiple Sclerosis.

**Figure S5.**
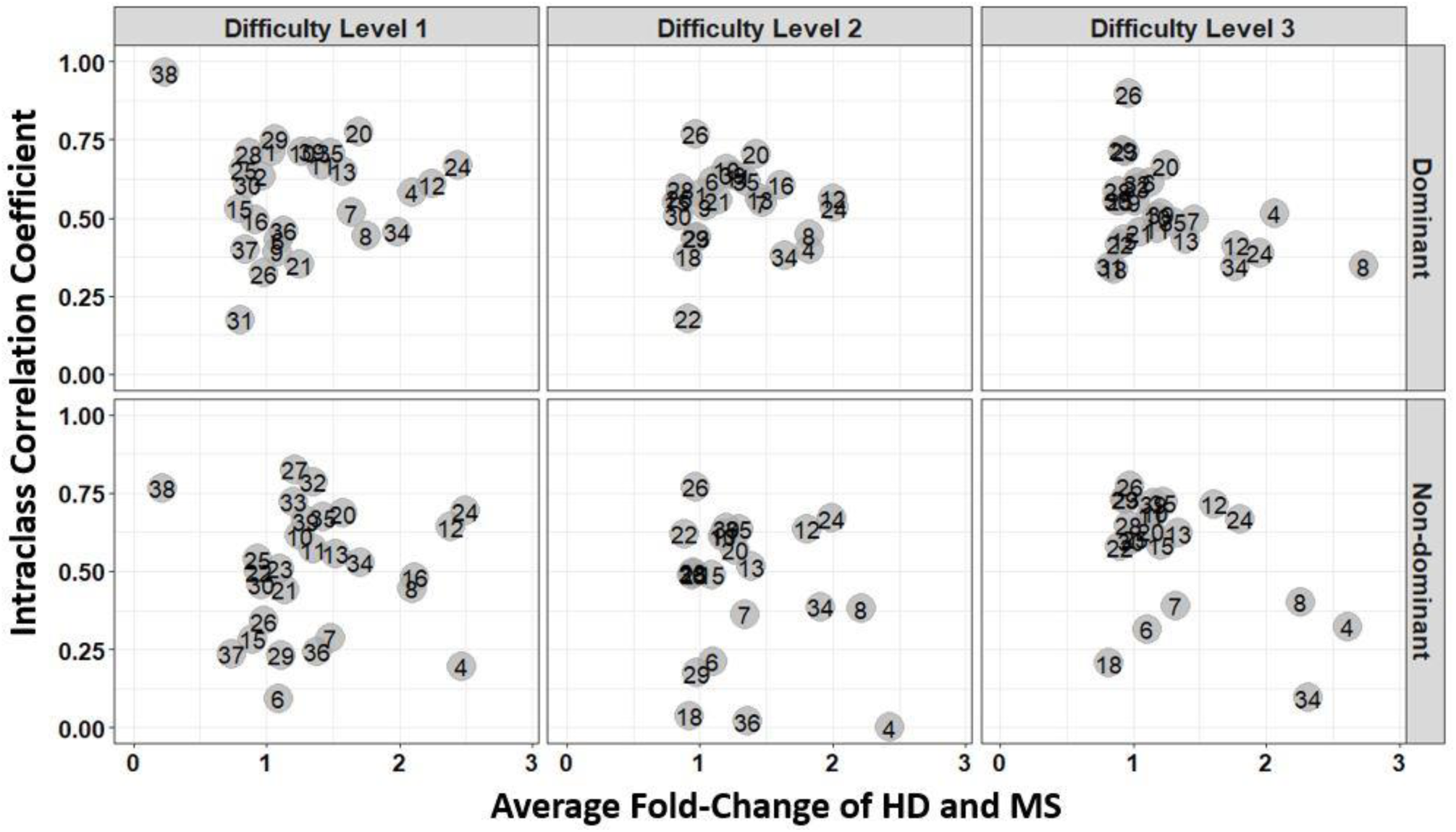
Average fold change of HD and MS of the spiral derived features with respect to their ICC from the training set. ICC was calculated from the granular data of the healthy donors (HD). The numbers indicate the feature’s labels as illustrated by the label in Table 2. HD and MS denote Healthy Donors and Multiple Sclerosis respectively.

**Figure S6.**
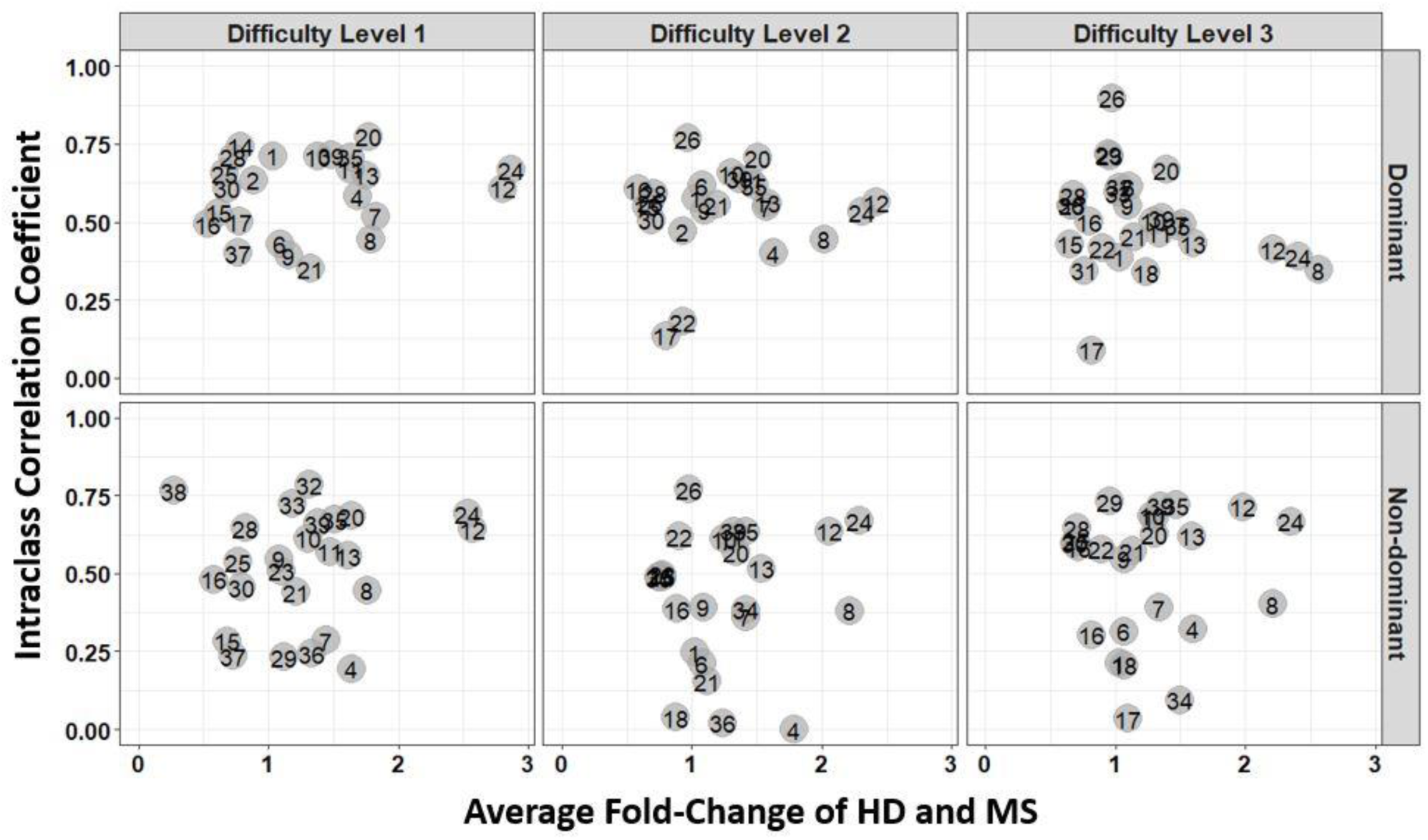
Average fold change of HD and MS of the spiral derived features with respect to their ICC from the test set. ICC was calculated from the granular data of the healthy donors (HD). The numbers illustrate the feature’s labels as indicated by the label in Table 2. HD and MS denote Healthy Donors and Multiple Sclerosis respectively.

**Figure S7.**
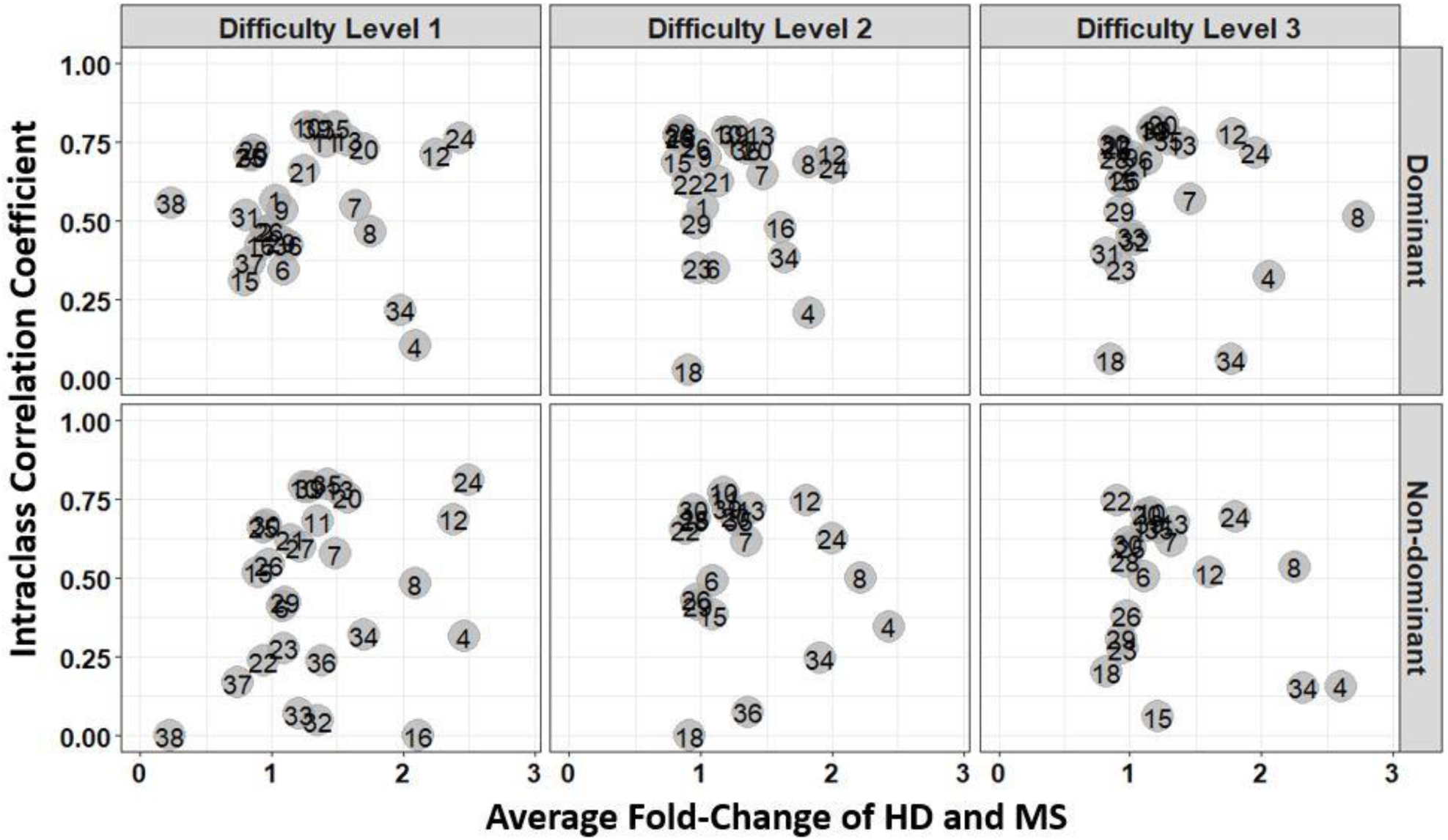
Average fold change of HD and MS of the spiral derived features with respect to their ICC from the training set. ICC was calculated from the granular data of the MS patients. The numbers indicate the feature’s labels as illustrated by the label in Table 2. HD and MS denote Healthy Donors and Multiple Sclerosis respectively.

**Figure S8.**
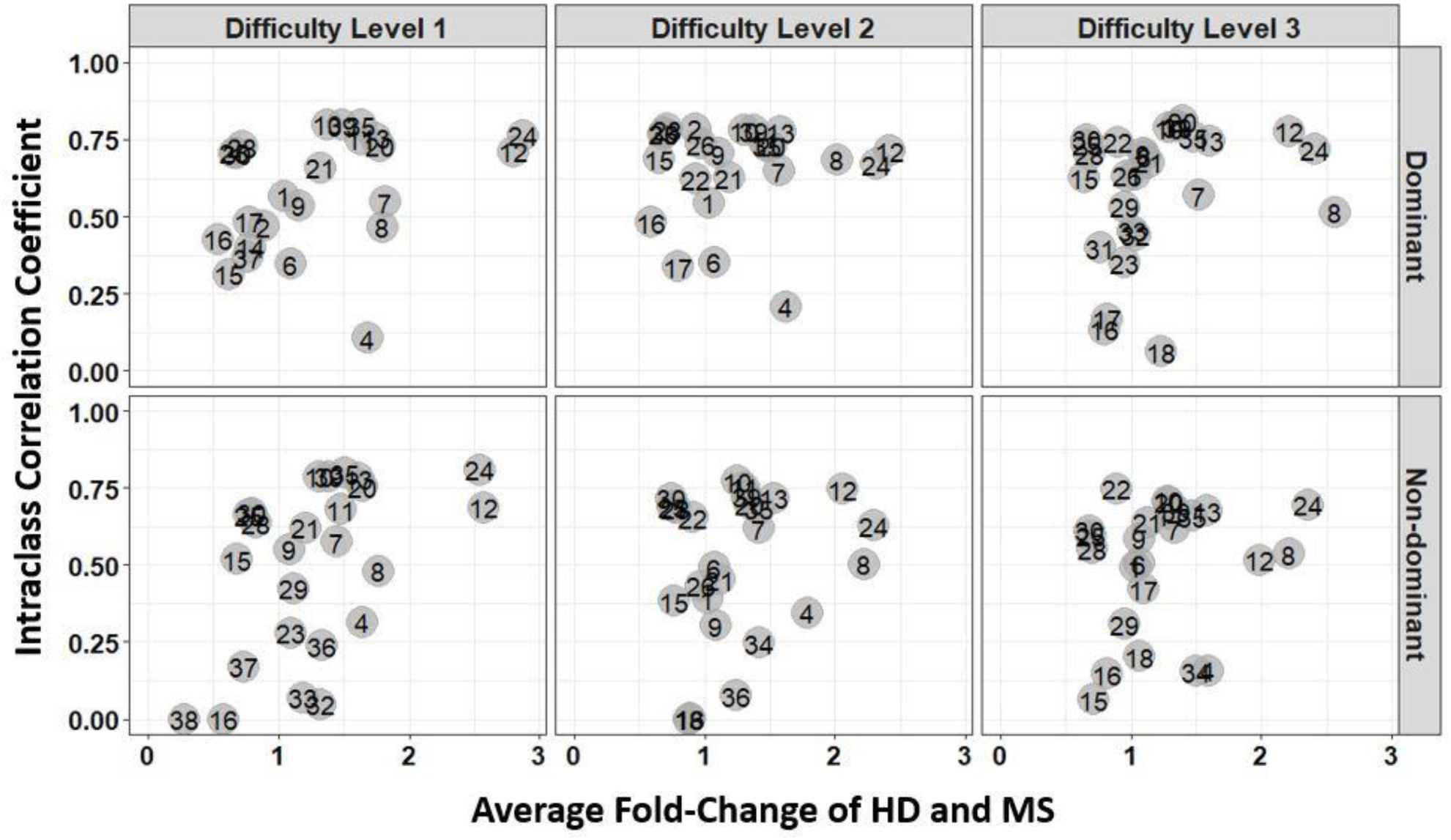
Average fold change of HD and MS of the spiral derived features with respect to their ICC from the test set. ICC was calculated from the granular data of the MS patients. The numbers illustrate the feature’s labels as indicated by the label in Table 2. HD and MS denote Healthy Donors and Multiple Sclerosis respectively.

**Figure S9.**
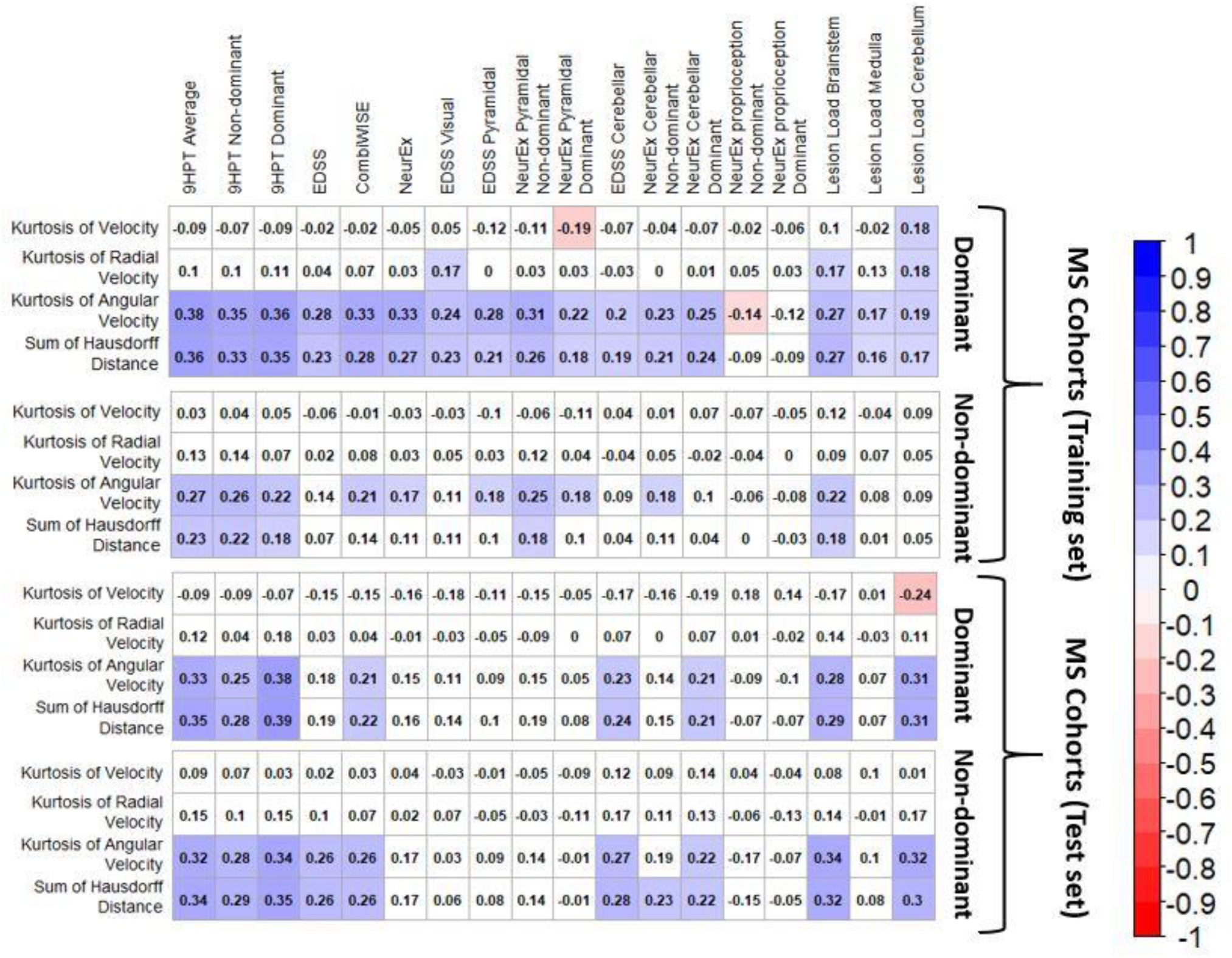
Spearman rho correlation matrix between the statistically significant clinical disability and the top four most significant spiral derived features based on FC at the difficulty level 3. The number indicates the spearman correlation coefficient. Red is negative correlation while blue stand for positive correlation. The white color indicates correlation that are not statistically significant at BH adjusted p-value of 0.05.

**Figure S10.**
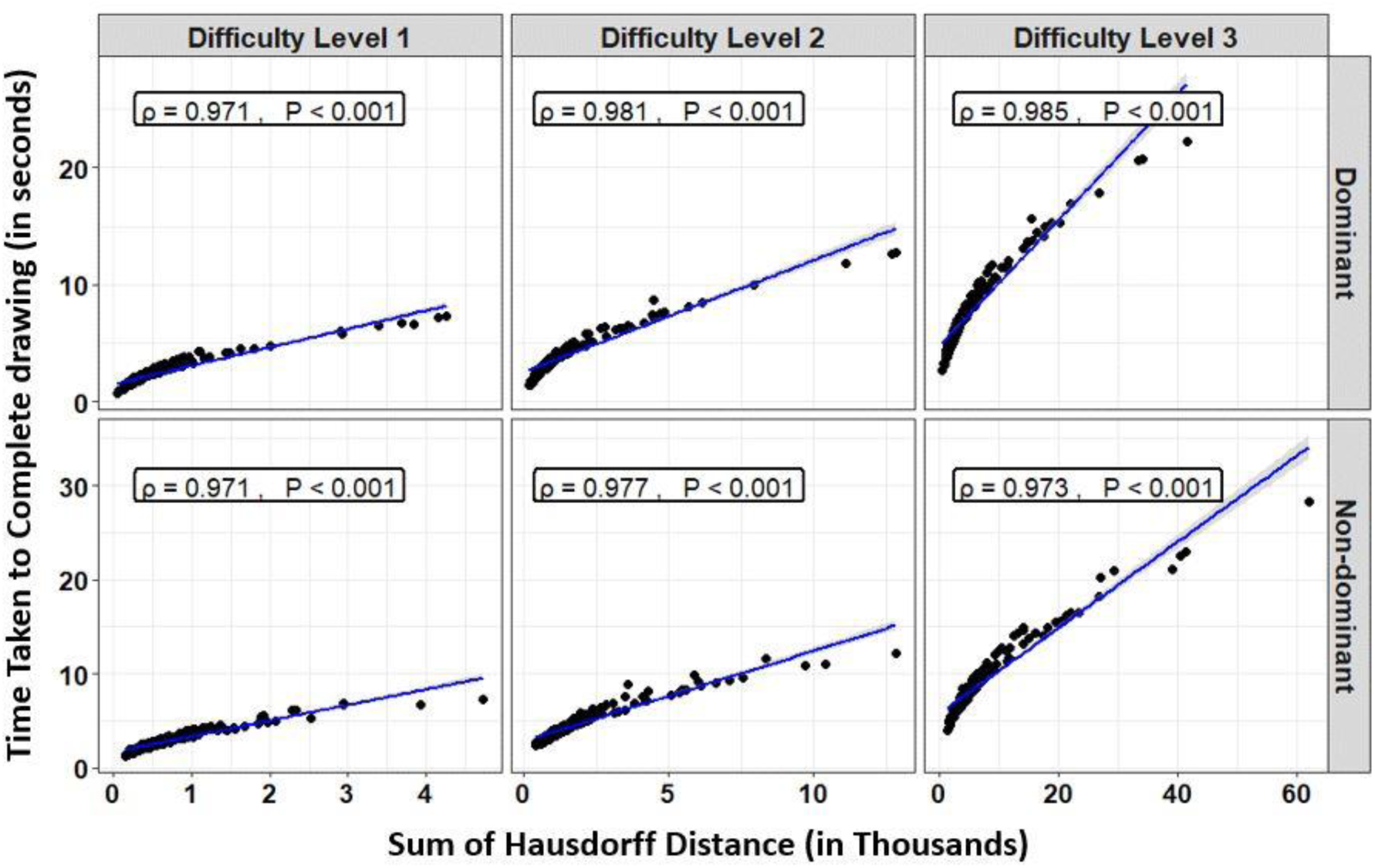
Relationship between the sum of the Hausdorff distances and the time taken to complete the spiral drawing among the healthy donors (HD; shown in black dots). Regression lines are shown in solid blue line. The ρ is the spearman correlation coefficient between the variables while P is the p-value of the correlation.

**Figure S11.**
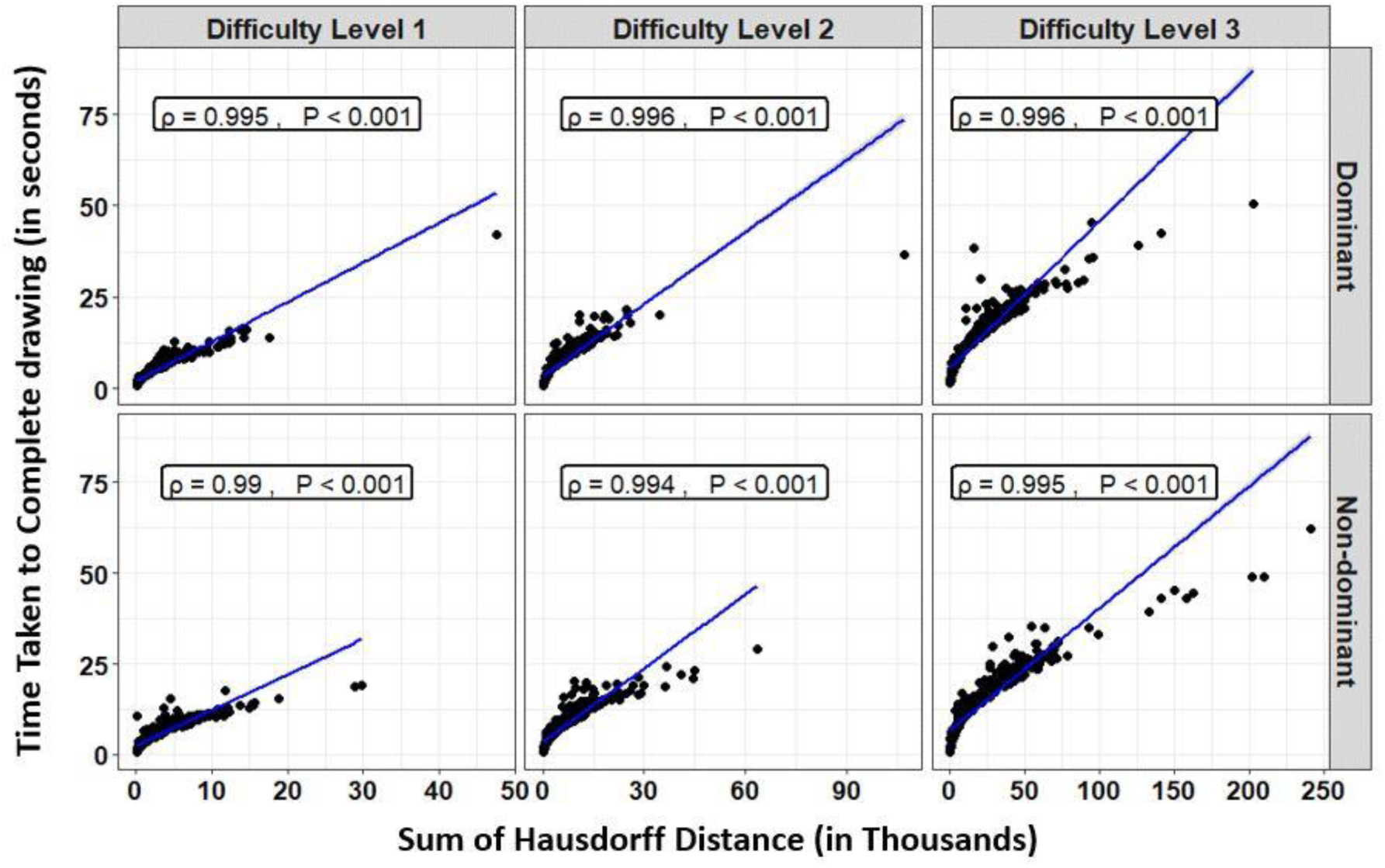
Relationship between the sum of the Hausdorff distances and the time taken to complete the spiral drawing among the MS patients – training set (shown in black dots). Regression lines are shown in solid blue line. The ρ is the spearman correlation coefficient between the variables while P is the p-value of the correlation. MS denotes Multiple Sclerosis.

**Figure S12.**
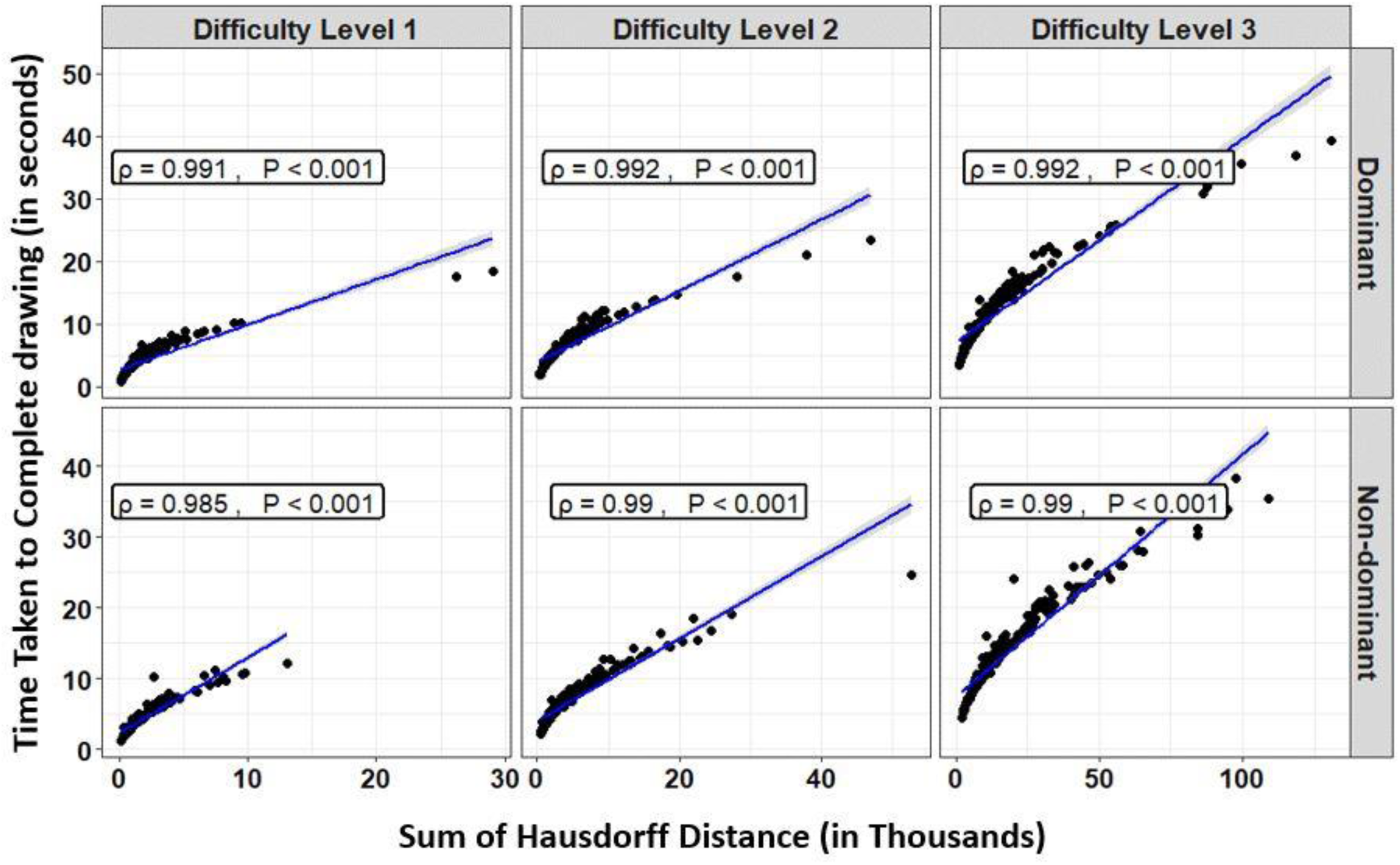
Relationship between the sum of the Hausdorff distances and the time taken to complete the spiral drawing among the MS patients – test set (shown in black dots). Regression lines are shown in solid blue line. The ρ is the spearman correlation coefficient between the variables while P is the p-value of the correlation. MS denotes Multiple Sclerosis.

**Figure S13.**
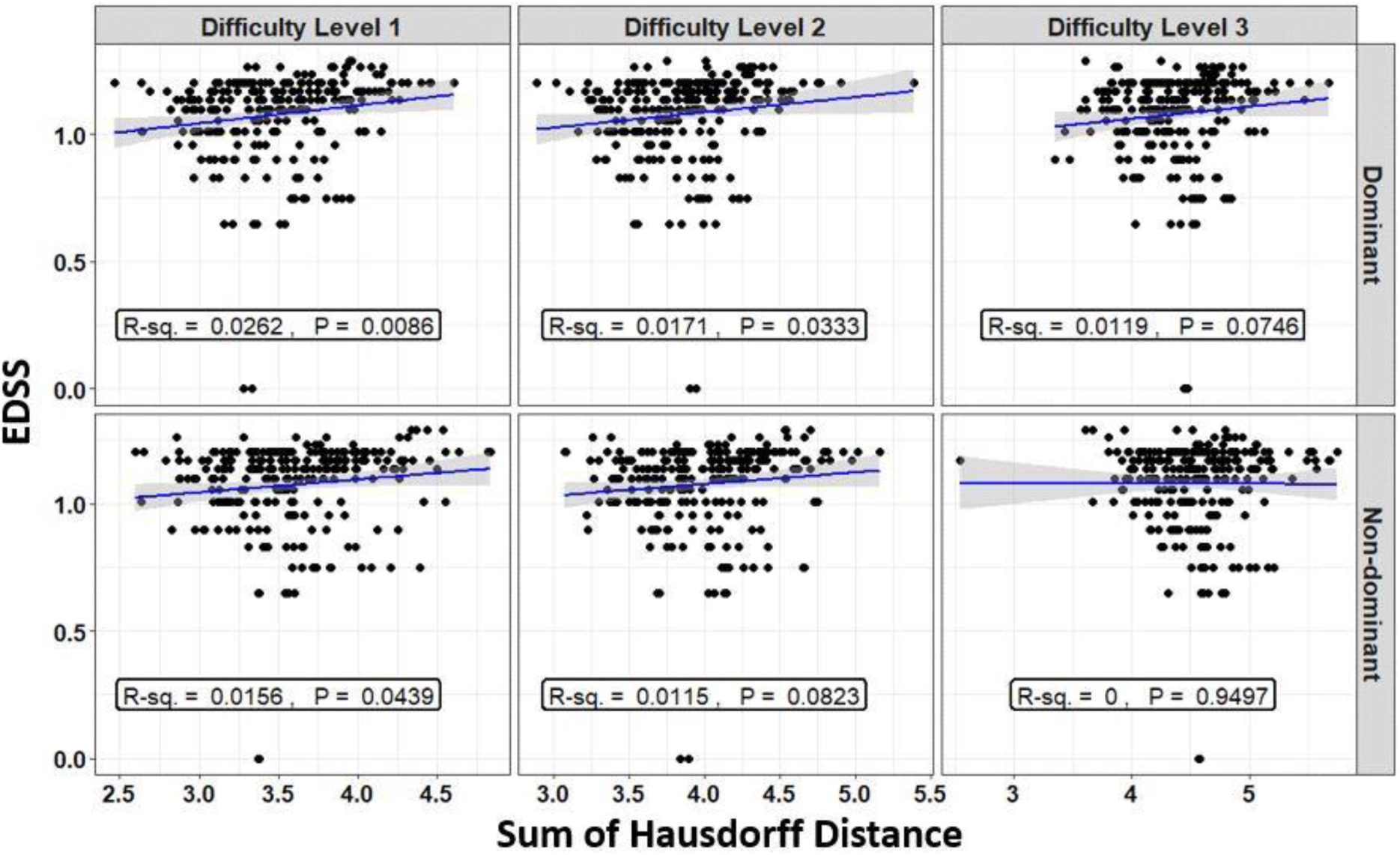
Relationship between the sum of the Hausdorff distances and EDSS of the MS patients in black dots. Linear regression lines are shown in solid blue line while the gray shaded area constitute the 95% confidence interval associated with the mean model’s prediction. The R-square denoted R-sq. indicates the percent of variance in EDSS that can be explained by the sum of Hausdorff distances. P is the model’s p-value. MS denotes Multiple Sclerosis.

**Figure S14.**
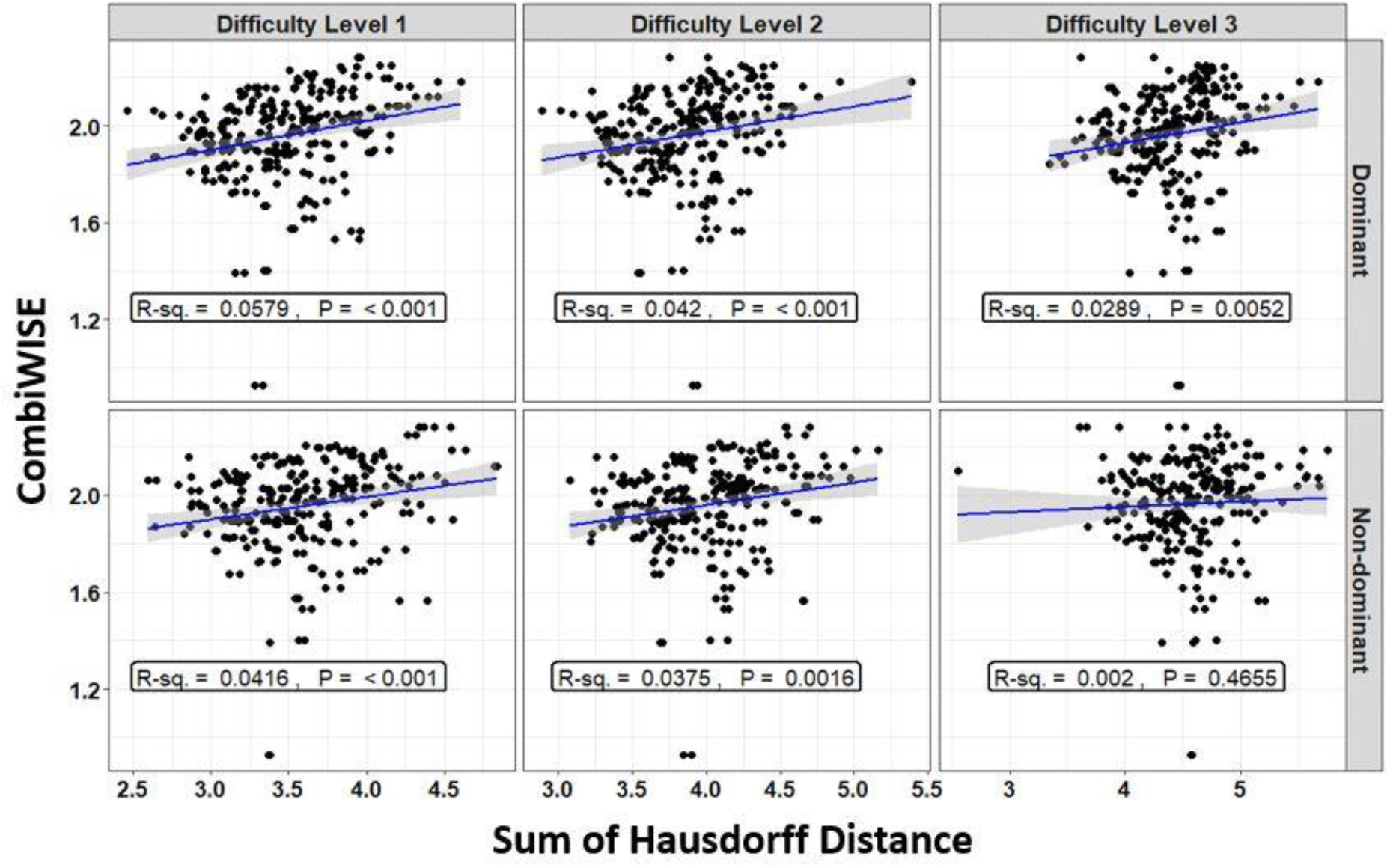
Relationship between the sum of the Hausdorff distances and CombiWISE of the MS patients in black dots. Linear regression lines are shown in solid blue line while the gray shaded area constitute the 95% confidence interval associated with the mean model’s prediction. The R-square denoted R-sq. indicates the percent of variance in CombiWISE that can be explained by the sum of Hausdorff distances. P is the model’s p-value. MS denotes Multiple Sclerosis.

**Figure S15.**
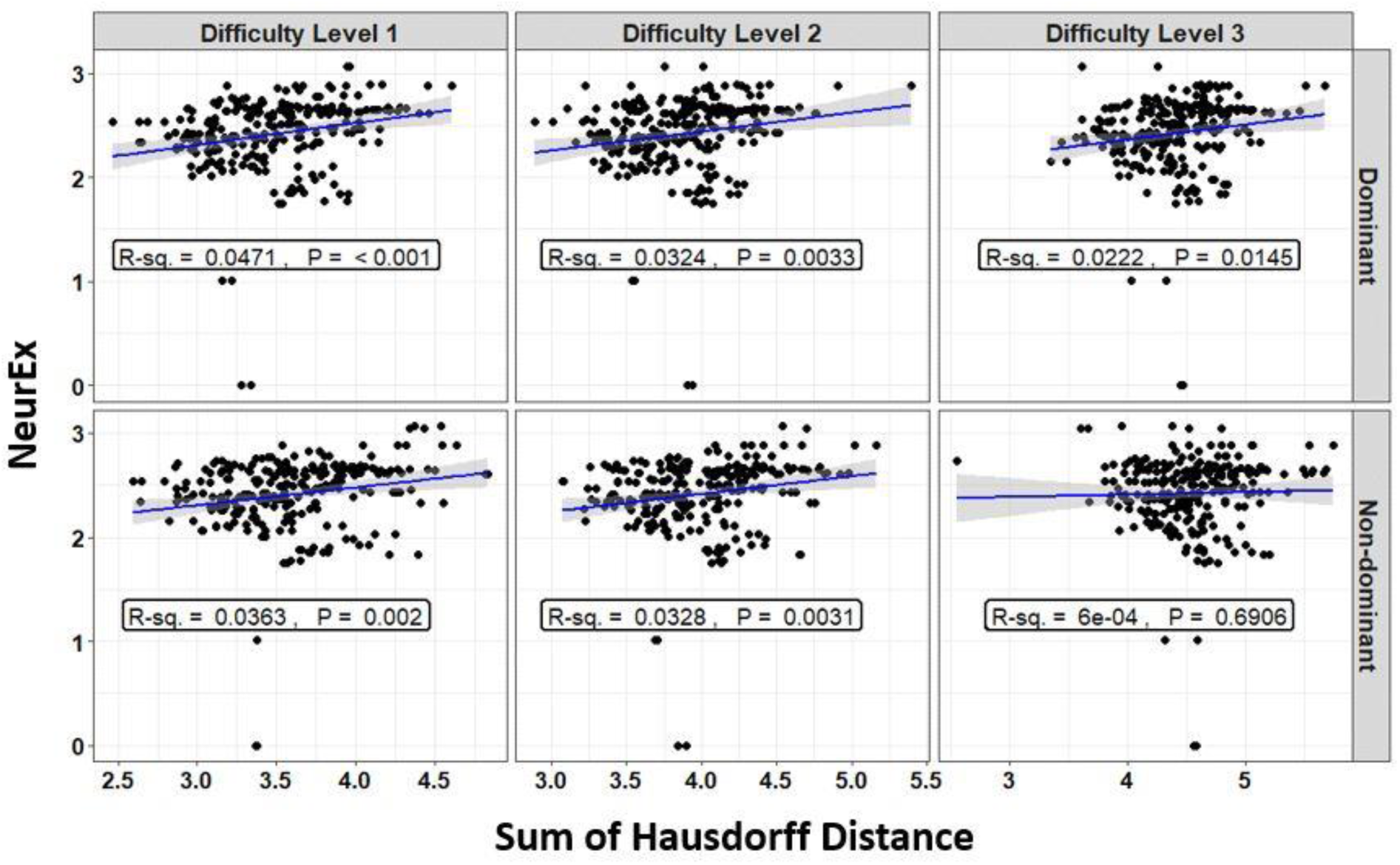
Relationship between the sum of the Hausdorff distances and NeurEx of the MS patients in black dots. Linear regression lines are shown in solid blue line while the gray shaded area constitute the 95% confidence interval associated with the mean model’s prediction. The R-square denoted R-sq. indicates the percent of variance in NeurEx that can be explained by the sum of Hausdorff distances. P is the model’s p-value. MS denotes Multiple Sclerosis.

**Table S1.** List of features relating to upper-extremity function calculated from the spiral drawing test. When applicable, references are provided for each feature calculated. Dashed symbols the references indicate not applicable.

| Label | Description | References |
| --- | --- | --- |
| F1 | Sum of velocity | - |
| F2 | Coefficient of variation of velocity | - |
| F3 | Skewness of velocity | - |
| F4 | Kurtosis of velocity | - |
| F5 | Sum of radial velocity | - |
| F6 | Coefficient of variation of radial velocity | - |
| F7 | Skewness of radial velocity | - |
| F8 | Kurtosis of radial velocity | - |
| F9 | Sum of angular velocity | - |
| F10 | Coefficient of variation of angular velocity | - |
| F11 | Skewness of angular velocity | - |
| F12 | Kurtosis of angular velocity | - |
| F13 | Sum of estimated pressure | - |
| F14 | Maximum Power Spectral Density (PSD) of velocity | (A. P. Creagh et al., 2020; Erasmus et al., 2001) |
| F15 | Dominant Frequency of velocity | (A. P. Creagh et al., 2020) |
| F16 | Maximum PSD of radial velocity | (A. P. Creagh et al., 2020; Erasmus et al., 2001) |
| F17 | Dominant Frequency of radial velocity | (A. P. Creagh et al., 2020) |
| F18 | Maximum PSD of angular velocity | (A. P. Creagh et al., 2020; Erasmus et al., 2001) |
| F19 | Dominant Frequency of angular velocity | (A. P. Creagh et al., 2020) |
| F20 | Approximate Entropy of velocity | (A. P. Creagh et al., 2020; Mevludin Memedi, Aghanavesi, & Westin, 2016; M. Memedi et al., 2015) |
| F21 | Approximate Entropy of radial velocity | (A. P. Creagh et al., 2020) |
| F22 | Approximate Entropy of angular velocity | (A. P. Creagh et al., 2020) |
| F23 | Maximum Hausdorff Distance (HD) | (A. P. Creagh et al., 2020; Dubuisson & Jain, 1994; Huttenlocher et al., 1993) |
| F24 | Sum of HD | (A. P. Creagh et al., 2020) |
| F25 | Sum of HD divided by time taken to complete drawing | (A. P. Creagh et al., 2020) |
| F26 | Interquartile Range of Sum of HD | (A. P. Creagh et al., 2020) |
| F27 | Sum of HD normalized by touchpoints at the beginning | (A. P. Creagh et al., 2020) |
| F28 | Sum of HD normalized by touchpoints at the end | (A. P. Creagh et al., 2020) |
| F29 | Sum of HD in the middle 15-85% | (A. P. Creagh et al., 2020) |
| F30 | Sum of HD in the middle 15-85% normalized by time taken to complete drawing | (A. P. Creagh et al., 2020) |
| F31 | Error calculated using Area Under the Curve | (A. P. Creagh et al., 2020) |
| F32 | Mean Square Error | (Asamoah et al., 2018; A. P. Creagh et al., 2020) |
| F33 | Root Mean Square Error | (A. P. Creagh et al., 2020) |
| F34 | Center of Shoot | (A. P. Creagh et al., 2020) |
| F35 | Time taken to complete drawing | - |
| F36 | Total Asymmetry of patient drawing | (A. P. Creagh et al., 2020) |
| F37 | True Asymmetry in comparison with reference shape | - |
| F38 | 2D image Correlation between two images | (Aljanabi et al., 2018; A. P. Creagh et al., 2020) |
| F39 | Image entropy of shape drawn | (A. P. Creagh et al., 2020; Tsai, Lee, & Matsuyama, 2008) |
| F40 | Image entropy of shape drawn with respect to reference shape | (A. P. Creagh et al., 2020) |

**Table S2.**
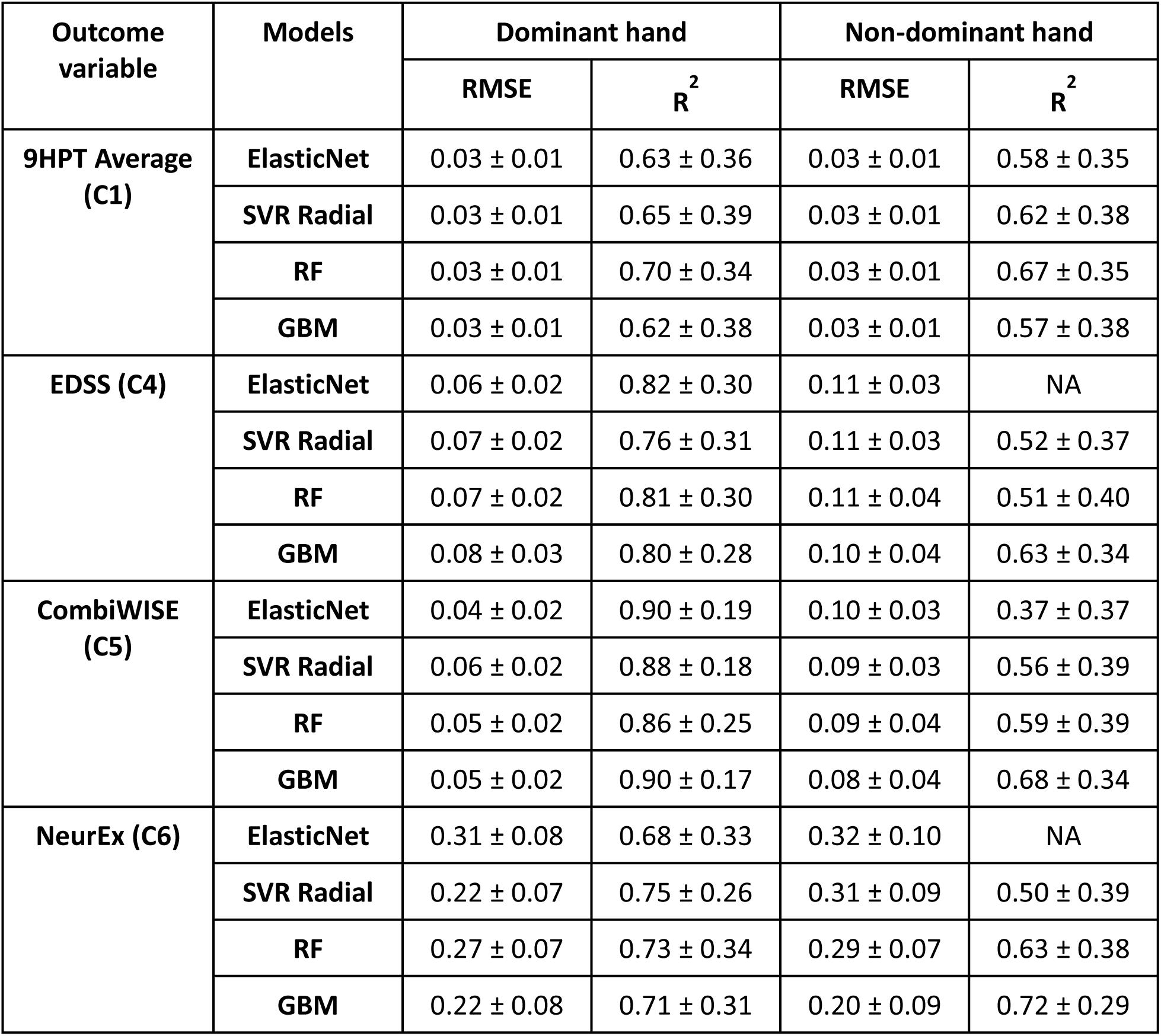
Root Mean Square Error (RMSE) and R-square (R^2^) of model predictions of the clinical disability scales per dominant and non-dominant hands among the healthy donors at the difficulty level 1. Models are build using 5-fold cross-validation (CV) with 10 repetitions. Results are provided using mean ± SD where the mean and SD are the mean and Standard Deviation across CV repetitions. ElasticNet, SVR Radial, RF, and GBM represent respectively the Elastic net, Support Vector Regression with Radial Basis Function kernel, Random Forest, and Stochastic Gradient Boosting regression models. NA indicates cannot be computed.

| Outcome variable | Models | Dominant hand |  | Non-dominant hand |  |
| --- | --- | --- | --- | --- | --- |
| | | RMSE | $R^2$ | RMSE | $R^2$ |
| 9HPT Average (C1) | ElasticNet | 0.03 ± 0.01 | 0.63 ± 0.36 | 0.03 ± 0.01 | 0.58 ± 0.35 |
|  | SVR Radial | 0.03 ± 0.01 | 0.65 ± 0.39 | 0.03 ± 0.01 | 0.62 ± 0.38 |
|  | RF | 0.03 ± 0.01 | 0.70 ± 0.34 | 0.03 ± 0.01 | 0.67 ± 0.35 |
|  | GBM | 0.03 ± 0.01 | 0.62 ± 0.38 | 0.03 ± 0.01 | 0.57 ± 0.38 |
| EDSS (C4) | ElasticNet | 0.06 ± 0.02 | 0.82 ± 0.30 | 0.11 ± 0.03 | NA |
|  | SVR Radial | 0.07 ± 0.02 | 0.76 ± 0.31 | 0.11 ± 0.03 | 0.52 ± 0.37 |
|  | RF | 0.07 ± 0.02 | 0.81 ± 0.30 | 0.11 ± 0.04 | 0.51 ± 0.40 |
|  | GBM | 0.08 ± 0.03 | 0.80 ± 0.28 | 0.10 ± 0.04 | 0.63 ± 0.34 |
| CombiWISE (C5) | ElasticNet | 0.04 ± 0.02 | 0.90 ± 0.19 | 0.10 ± 0.03 | 0.37 ± 0.37 |
|  | SVR Radial | 0.06 ± 0.02 | 0.88 ± 0.18 | 0.09 ± 0.03 | 0.56 ± 0.39 |
|  | RF | 0.05 ± 0.02 | 0.86 ± 0.25 | 0.09 ± 0.04 | 0.59 ± 0.39 |
|  | GBM | 0.05 ± 0.02 | 0.90 ± 0.17 | 0.08 ± 0.04 | 0.68 ± 0.34 |
| NeurEx (C6) | ElasticNet | 0.31 ± 0.08 | 0.68 ± 0.33 | 0.32 ± 0.10 | NA |
|  | SVR Radial | 0.22 ± 0.07 | 0.75 ± 0.26 | 0.31 ± 0.09 | 0.50 ± 0.39 |
|  | RF | 0.27 ± 0.07 | 0.73 ± 0.34 | 0.29 ± 0.07 | 0.63 ± 0.38 |
|  | GBM | 0.22 ± 0.08 | 0.71 ± 0.31 | 0.20 ± 0.09 | 0.72 ± 0.29 |

**Table S3.** Root Mean Square Error (RMSE) and R-square (R^2^) of model predictions of the clinical disability scales per dominant and non-dominant hands among the MS patients at the difficulty level 1. Models are build using 5-fold cross-validation (CV) with 10 repetitions. Results are provided using mean ± SD where the mean and SD are the mean and Standard Deviation across CV repetitions. ElasticNet, SVR Radial, RF, and GBM represent respectively the Elastic net, Support Vector Regression with Radial Basis Function kernel, Random Forest, and Stochastic Gradient Boosting regression models. NA indicate cannot be computed. MS denotes Multiple Sclerosis.

| Outcome variable | Models | Dominant hand |  | Non-dominant hand |  |
| --- | --- | --- | --- | --- | --- |
| | | RMSE | $R^2$ | RMSE | $R^2$ |
| 9HPT Average (C1) | ElasticNet | $0.29 \pm 0.04$ | $0.21 \pm 0.05$ | $0.30 \pm 0.05$ | $0.19 \pm 0.07$ |
| | SVR Radial | $0.29 \pm 0.07$ | $0.23 \pm 0.11$ | $0.31 \pm 0.05$ | $0.19 \pm 0.10$ |
| | RF | $0.29 \pm 0.03$ | $0.21 \pm 0.10$ | $0.31 \pm 0.04$ | $0.16 \pm 0.08$ |
| | GBM | $0.30 \pm 0.04$ | $0.19 \pm 0.09$ | $0.31 \pm 0.04$ | $0.15 \pm 0.08$ |
| EDSS (C4) | ElasticNet | $0.16 \pm 0.03$ | $0.07 \pm 0.05$ | $0.17 \pm 0.03$ | NA |
| | SVR Radial | $0.17 \pm 0.03$ | $0.06 \pm 0.05$ | $0.17 \pm 0.04$ | $0.04 \pm 0.04$ |
| | RF | $0.17 \pm 0.03$ | $0.07 \pm 0.06$ | $0.17 \pm 0.03$ | $0.03 \pm 0.03$ |
| | GBM | $0.17 \pm 0.03$ | $0.06 \pm 0.05$ | $0.17 \pm 0.04$ | $0.02 \pm 0.02$ |
| CombiWISE (C5) | ElasticNet | $0.18 \pm 0.03$ | $0.10 \pm 0.05$ | $0.18 \pm 0.03$ | $0.05 \pm 0.05$ |
| | SVR Radial | $0.18 \pm 0.03$ | $0.12 \pm 0.07$ | $0.18 \pm 0.03$ | $0.08 \pm 0.07$ |
| | RF | $0.18 \pm 0.03$ | $0.10 \pm 0.06$ | $0.18 \pm 0.03$ | $0.06 \pm 0.05$ |
| | GBM | $0.18 \pm 0.03$ | $0.10 \pm 0.06$ | $0.19 \pm 0.03$ | $0.05 \pm 0.05$ |
| NeurEx (C6) | ElasticNet | $0.35 \pm 0.08$ | $0.10 \pm 0.06$ | $0.36 \pm 0.08$ | $0.04 \pm 0.04$ |
| | SVR Radial | $0.35 \pm 0.08$ | $0.12 \pm 0.06$ | $0.36 \pm 0.09$ | $0.06 \pm 0.05$ |
| | RF | $0.35 \pm 0.08$ | $0.11 \pm 0.07$ | $0.37 \pm 0.08$ | $0.06 \pm 0.06$ |
| | GBM | $0.36 \pm 0.08$ | $0.09 \pm 0.05$ | $0.36 \pm 0.08$ | $0.05 \pm 0.04$ |

**Table S4.**
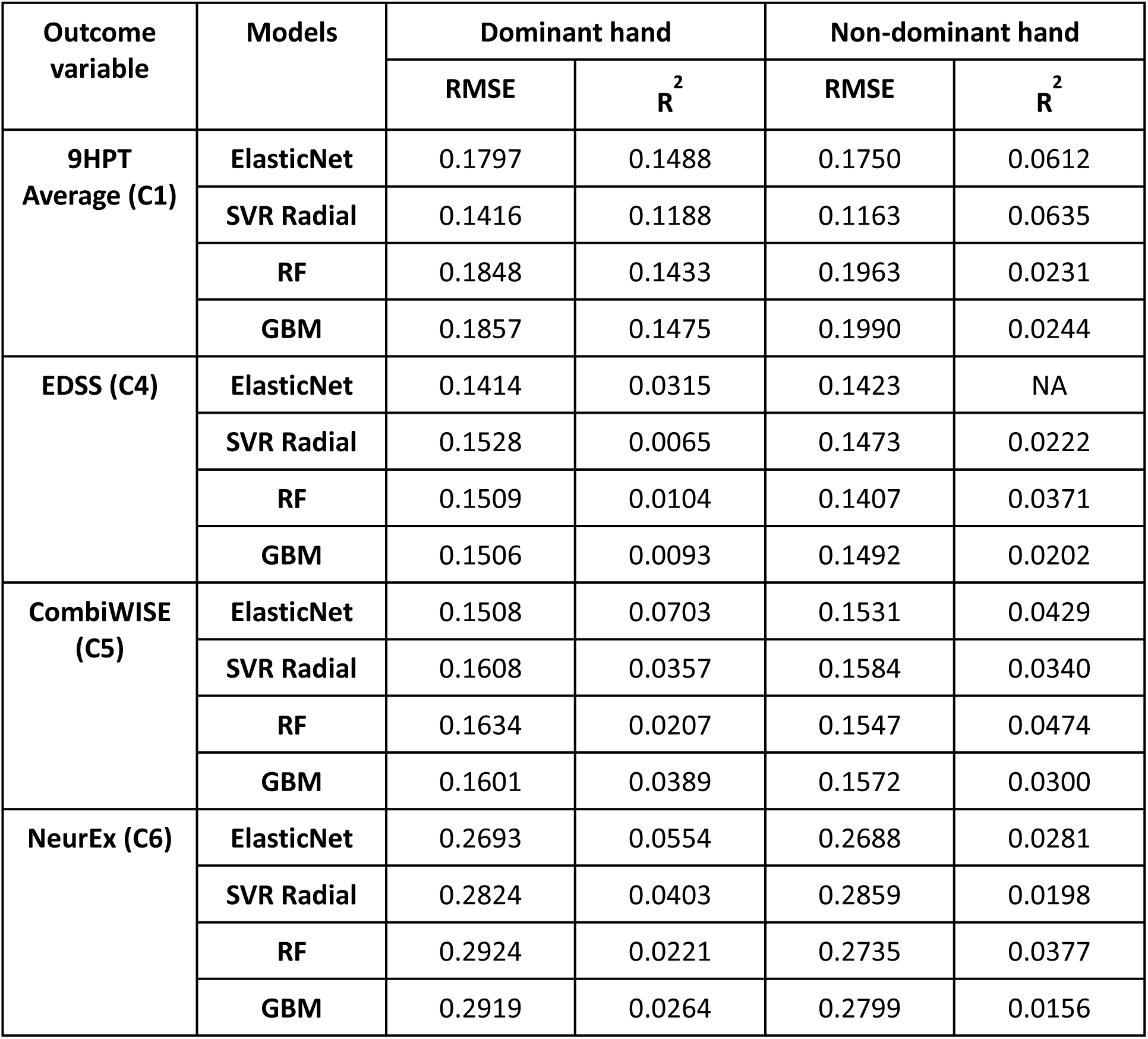
The out-of-sample test performance of the clinical disability scales versus the top four most significant spiral derived features (Kurtosis of velocity, radial velocity, angular velocity, and the sum of Hausdorff Distances). The test performance was measured using Root Mean Square Error (RMSE) and R-square (R^2^) of model predictions per dominant and non-dominant hands among the MS cohorts at the difficulty level 1. ElasticNet, SVR Radial, RF, and GBM represent respectively the Elastic net, Support Vector Regression with Radial Basis Function kernel, Random Forest, and Stochastic Gradient Boosting regression models. NA indicate cannot be computed.

| Outcome variable | Models | Dominant hand |  | Non-dominant hand |  |
| --- | --- | --- | --- | --- | --- |
| | | RMSE | $R^2$ | RMSE | $R^2$ |
| 9HPT Average (C1) | ElasticNet | 0.1797 | 0.1488 | 0.1750 | 0.0612 |
|  | SVR Radial | 0.1416 | 0.1188 | 0.1163 | 0.0635 |
|  | RF | 0.1848 | 0.1433 | 0.1963 | 0.0231 |
|  | GBM | 0.1857 | 0.1475 | 0.1990 | 0.0244 |
| EDSS (C4) | ElasticNet | 0.1414 | 0.0315 | 0.1423 | NA |
|  | SVR Radial | 0.1528 | 0.0065 | 0.1473 | 0.0222 |
|  | RF | 0.1509 | 0.0104 | 0.1407 | 0.0371 |
|  | GBM | 0.1506 | 0.0093 | 0.1492 | 0.0202 |
| CombiWISE (C5) | ElasticNet | 0.1508 | 0.0703 | 0.1531 | 0.0429 |
|  | SVR Radial | 0.1608 | 0.0357 | 0.1584 | 0.0340 |
|  | RF | 0.1634 | 0.0207 | 0.1547 | 0.0474 |
|  | GBM | 0.1601 | 0.0389 | 0.1572 | 0.0300 |
| NeurEx (C6) | ElasticNet | 0.2693 | 0.0554 | 0.2688 | 0.0281 |
|  | SVR Radial | 0.2824 | 0.0403 | 0.2859 | 0.0198 |
|  | RF | 0.2924 | 0.0221 | 0.2735 | 0.0377 |
|  | GBM | 0.2919 | 0.0264 | 0.2799 | 0.0156 |

**Table S5.**
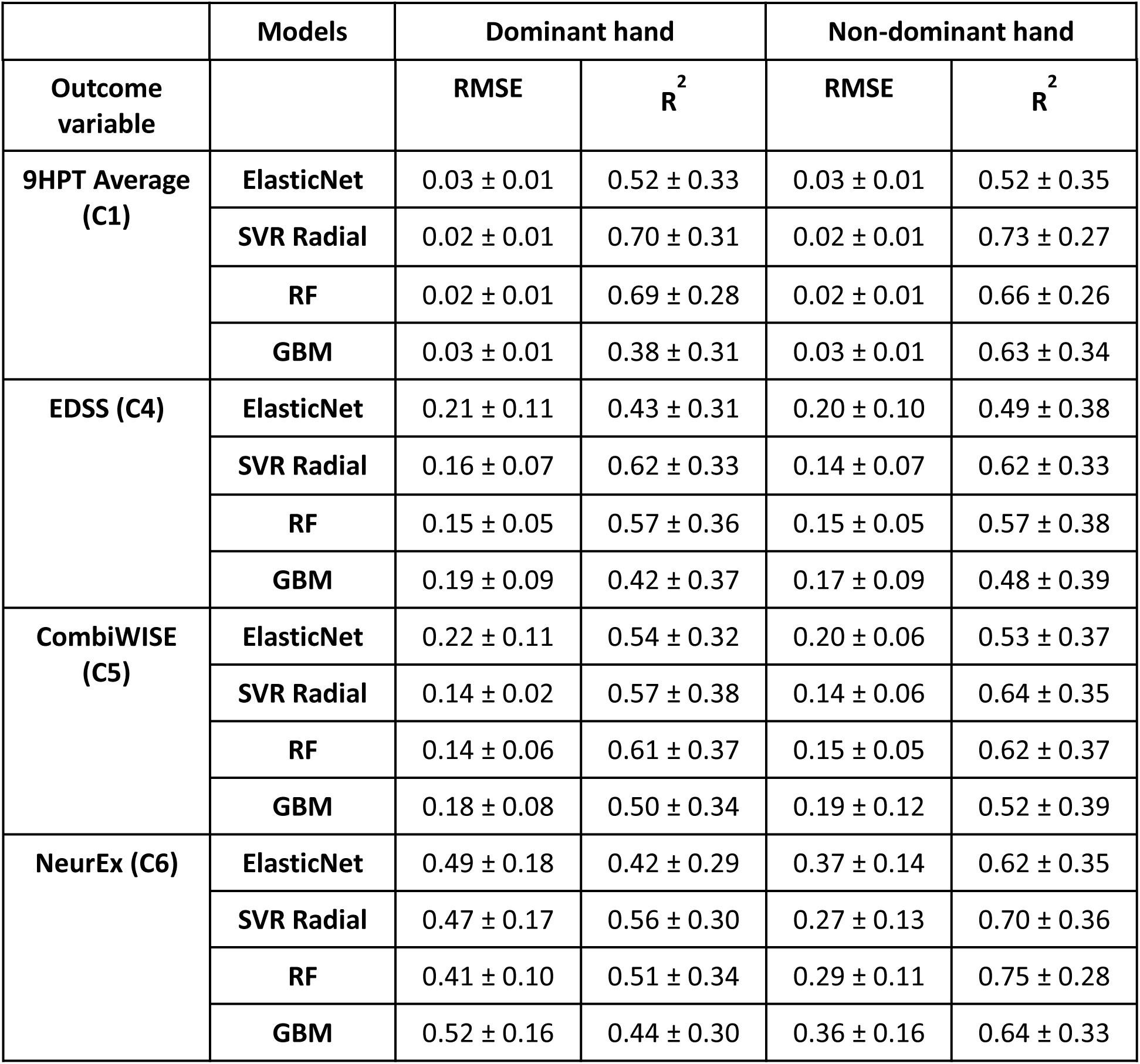
Root Mean Square Error (RMSE) and R-square (R^2^) of model predictions of the clinical disability scales per dominant and non-dominant hands among the healthy donors (HD) at the difficulty level 2. Models are build using 5-fold cross-validation (CV) with 10 repetitions. Results are provided using mean ± SD where the mean and SD are the mean and Standard Deviation across CV repetitions. ElasticNet, SVR Radial, RF, and GBM represent respectively the Elastic net, Support Vector Regression with Radial Basis Function kernel, Random Forest, and Stochastic Gradient Boosting regression models.

**Table S6.**
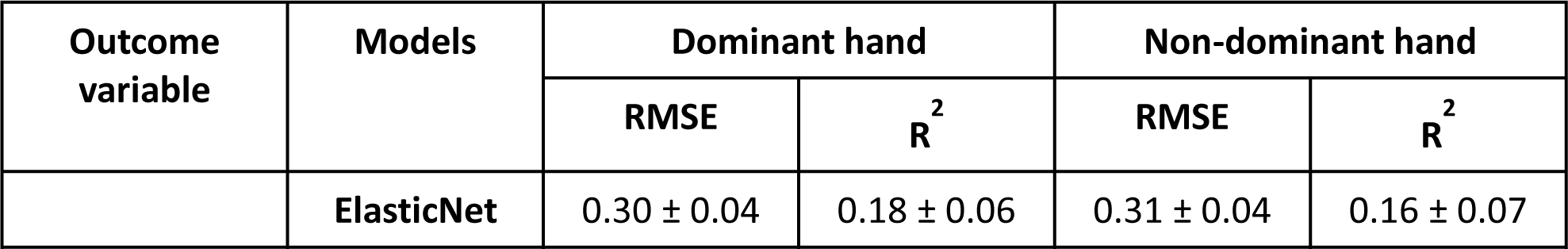
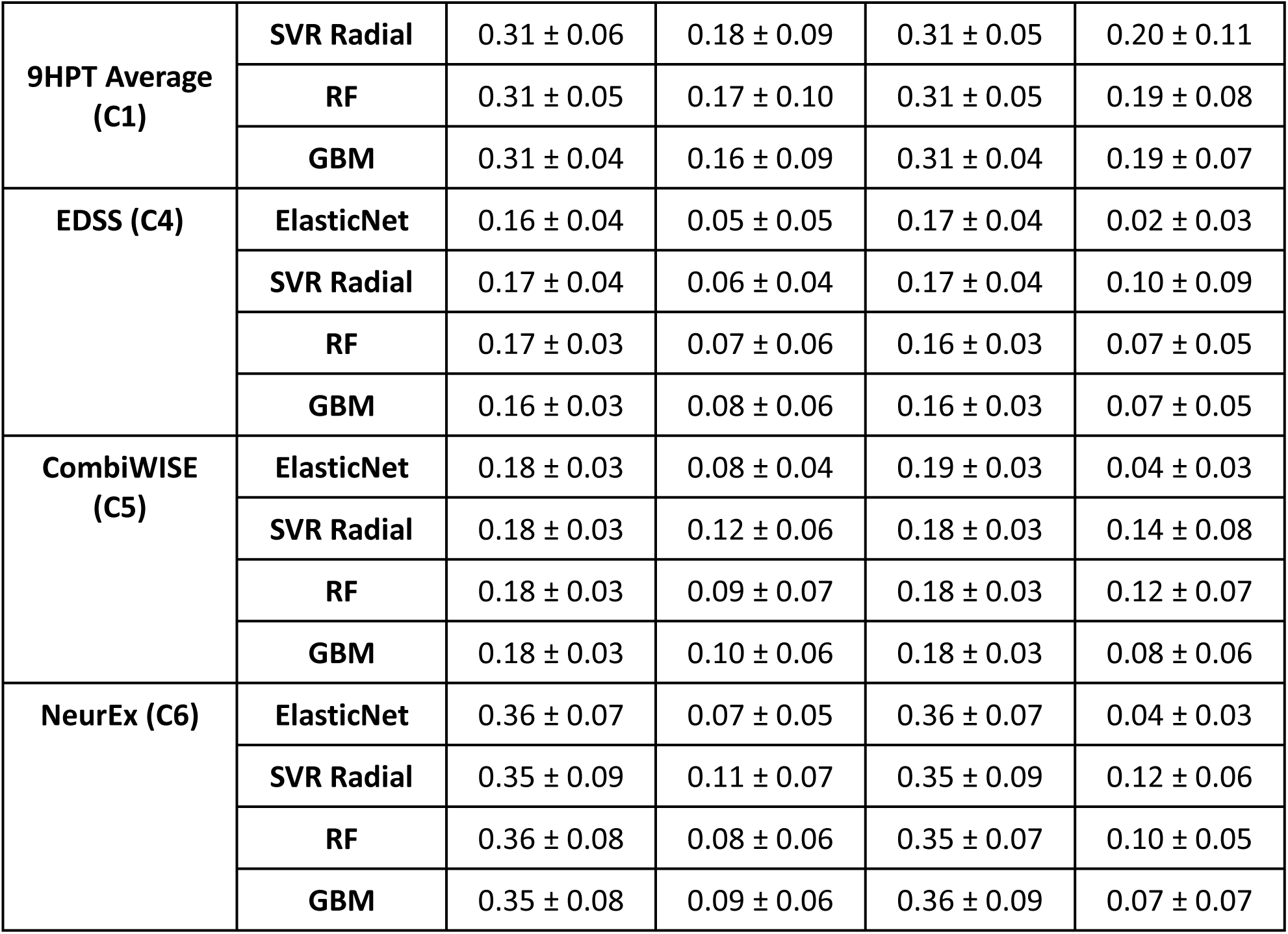
Root Mean Square Error (RMSE) and R-square (R^2^) of model predictions of the clinical disability scales per dominant and non-dominant hands among the MS patients at the difficulty level 2. Models are build using 5-fold cross-validation (CV) with 10 repetitions. Results are provided using mean ± SD where the mean and SD are the mean and Standard Deviation across CV repetitions. ElasticNet, SVR Radial, RF, and GBM represent respectively the Elastic net, Support Vector Regression with Radial Basis Function kernel, Random Forest, and Stochastic Gradient Boosting regression models. MS denotes Multiple Sclerosis.

**Table S7.** The out-of-sample test performance of the clinical disability scales versus the top four most significant spiral derived features (Kurtosis of velocity, radial velocity, angular velocity, and the sum of Hausdorff Distances). The test performance was measured using Root Mean Square Error (RMSE) and R-square (R^2^) of model predictions per dominant and non-dominant hands among the MS cohorts at the difficulty level 2. ElasticNet, SVR Radial, RF, and GBM represent respectively the Elastic net, Support Vector Regression with Radial Basis Function kernel, Random Forest, and Stochastic Gradient Boosting regression models.

| **Outcome variable** | **Models** | **Dominant hand** |  | **Non-dominant hand** |  |
| --- | --- | --- | --- | --- | --- |
|  |  | **RMSE** | **$R^2$** | **RMSE** | **$R^2$** |
| **9HPT Average (C1)** | **ElasticNet** | 0.1683 | 0.1813 | 0.1789 | 0.0632 |
|  | **SVR Radial** | 0.1292 | 0.1666 | 0.1677 | 0.0079 |
|  | **RF** | 0.1870 | 0.1751 | 0.1945 | 0.0537 |
|  | **GBM** | 0.1867 | 0.1590 | 0.2049 | 0.0564 |
| **EDSS (C4)** | **ElasticNet** | 0.1414 | 0.0195 | 0.1409 | 0.0364 |
|  | **SVR Radial** | 0.1560 | 0.0155 | 0.1505 | 0.0032 |
|  | **RF** | 0.1469 | 0.0317 | 0.1479 | 0.0098 |
|  | **GBM** | 0.1442 | 0.0395 | 0.1511 | 0.0065 |
| **CombiWISE (C5)** | **ElasticNet** | 0.1528 | 0.0526 | 0.1533 | 0.0444 |
|  | **SVR Radial** | 0.1601 | 0.0482 | 0.1658 | 0.0017 |
|  | **RF** | 0.1580 | 0.0561 | 0.1622 | 0.0152 |
|  | **GBM** | 0.1577 | 0.0622 | 0.1653 | 0.0233 |
| **NeurEx (C6)** | **ElasticNet** | 0.2692 | 0.0432 | 0.2703 | 0.0251 |
|  | **SVR Radial** | 0.2811 | 0.0467 | 0.2975 | 0.0002 |
|  | **RF** | 0.2809 | 0.0481 | 0.2849 | 0.0200 |
|  | **GBM** | 0.2779 | 0.0643 | 0.2822 | 0.0184 |

**Table S8.** Root Mean Square Error (RMSE) and R-square (R^2^) of model predictions of the clinical disability scales per dominant and non-dominant hands among the healthy donors (HD) at the difficulty level 3. Models are build using 5-fold cross-validation (CV) with 10 repetitions. Results are provided using mean ± SD where the mean and SD are the mean and Standard Deviation across CV repetitions. ElasticNet, SVR Radial, RF, and GBM represent respectively the Elastic net, Support Vector Regression with Radial Basis Function kernel, Random Forest, and Stochastic Gradient Boosting regression models. NA indicates cannot be computed.

| **Outcome variable** | **Models** | **Dominant hand** |  | **Non-dominant hand** |  |
| --- | --- | --- | --- | --- | --- |
|  |  | **RMSE** | **$R^2$** | **RMSE** | **$R^2$** |
| **9HPT Average (C1)** | **ElasticNet** | 0.03 ± 0.01 | 0.43 ± 0.34 | 0.02 ± 0.01 | 0.66 ± 0.33 |
| | **SVR Radial** | 0.03 ± 0.01 | 0.49 ± 0.36 | 0.02 ± 0.01 | 0.59 ± 0.35 |
| | **RF** | 0.03 ± 0.01 | 0.56 ± 0.34 | 0.03 ± 0.01 | 0.73 ± 0.28 |
| | **GBM** | 0.03 ± 0.01 | 0.54 ± 0.35 | 0.03 ± 0.01 | 0.66 ± 0.28 |
| **EDSS (C4)** | **ElasticNet** | 0.22 ± 0.12 | NA | 0.24 ± 0.12 | NA |
| | **SVR Radial** | 0.21 ± 0.14 | 0.35 ± 0.32 | 0.21 ± 0.13 | 0.54 ± 0.34 |
| | **RF** | 0.26 ± 0.12 | 0.30 ± 0.27 | 0.27 ± 0.12 | 0.45 ± 0.37 |
|  | **GBM** | 0.28 ± 0.13 | 0.37 ± 0.36 | 0.31 ± 0.13 | 0.41 ± 0.33 |
| **CombiWISE (C5)** | **ElasticNet** | 0.23 ± 0.13 | NA | 0.23 ± 0.13 | NA |
|  | **SVR Radial** | 0.21 ± 0.14 | 0.47 ± 0.36 | 0.20 ± 0.16 | 0.53 ± 0.36 |
|  | **RF** | 0.26 ± 0.12 | 0.36 ± 0.29 | 0.27 ± 0.16 | 0.50 ± 0.30 |
|  | **GBM** | 0.29 ± 0.12 | 0.40 ± 0.31 | 0.30 ± 0.15 | 0.47 ± 0.30 |
| **NeurEx (C6)** | **ElasticNet** | 0.53 ± 0.22 | 0.42 ± 0.34 | 0.57 ± 0.24 | NA |
|  | **SVR Radial** | 0.52 ± 0.29 | 0.30 ± 0.33 | 0.50 ± 0.24 | 0.60 ± 0.35 |
|  | **RF** | 0.59 ± 0.22 | 0.30 ± 0.31 | 0.62 ± 0.25 | 0.44 ± 0.35 |
|  | **GBM** | 0.66 ± 0.31 | 0.43 ± 0.33 | 0.69 ± 0.25 | 0.52 ± 0.38 |

**Table S9.**
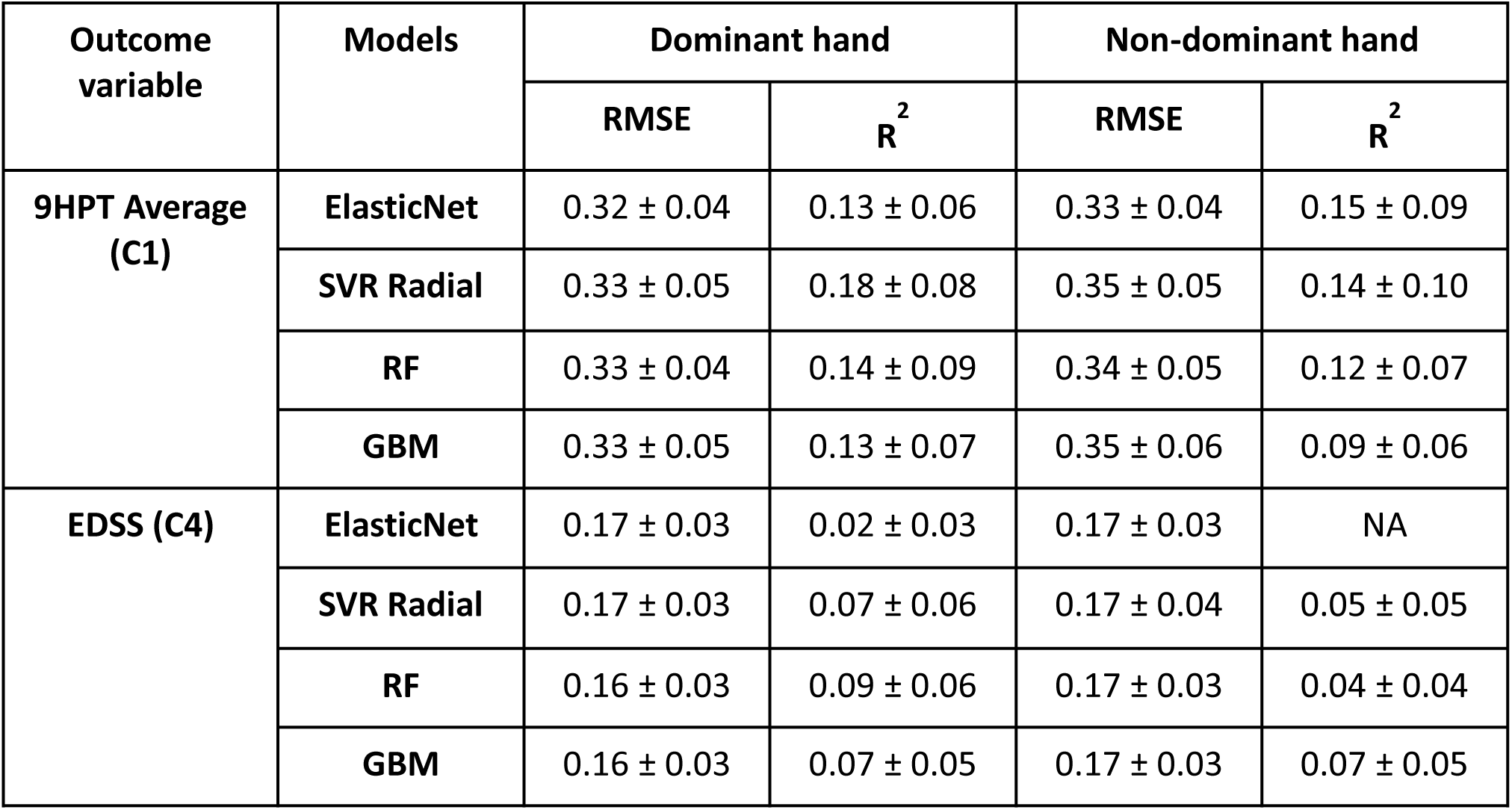
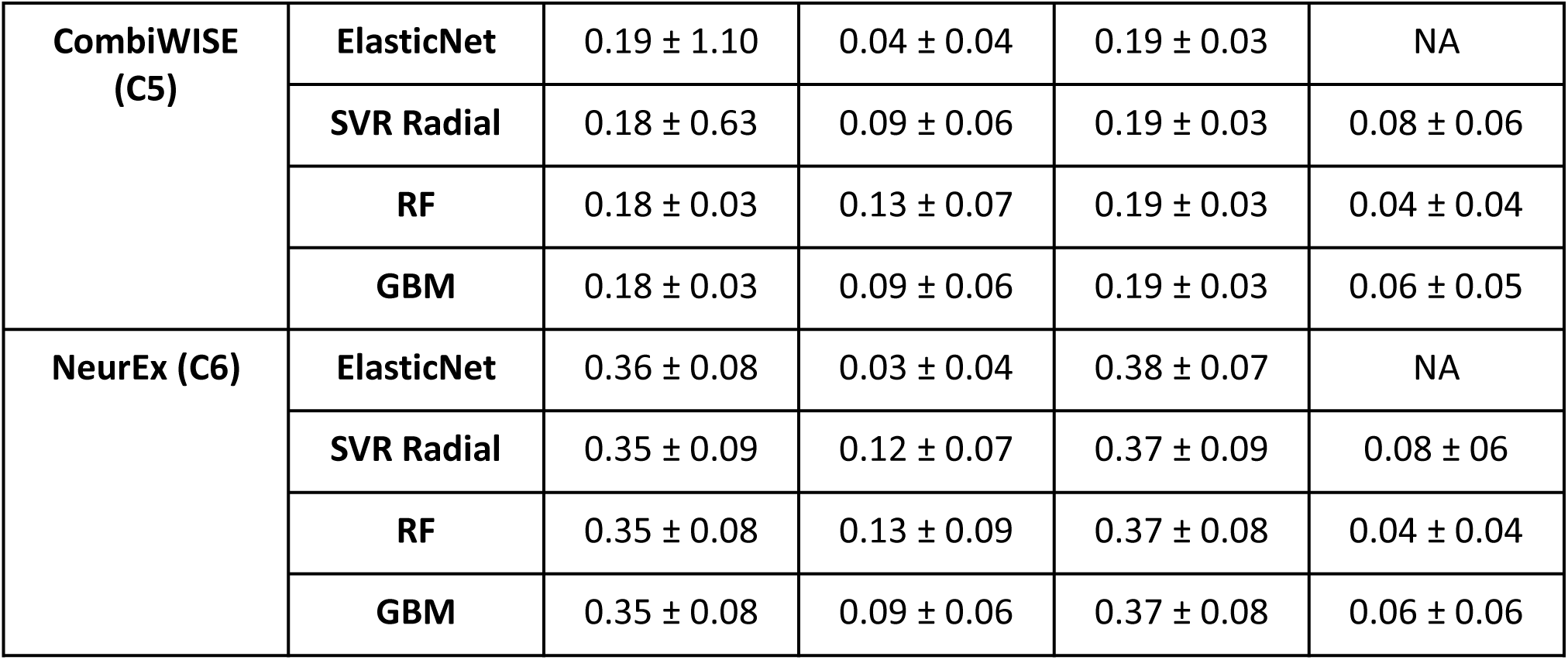
Root Mean Square Error (RMSE) and R-square (R^2^) of model predictions of the clinical disability scales per dominant and non-dominant hands among the MS patients at the difficulty level 2. Models are build using 5-fold cross-validation (CV) with 10 repetitions. Results are provided using mean ± SD where the mean and SD are the mean and Standard Deviation across CV repetitions. ElasticNet, SVR Radial, RF, and GBM represent respectively the Elastic net, Support Vector Regression with Radial Basis Function kernel, Random Forest, and Stochastic Gradient Boosting regression models. NA indicates cannot be computed. MS denotes Multiple Sclerosis.

**Table S10.** The out-of-sample test performance of the clinical disability scales versus the top four most significant spiral derived features (Kurtosis of velocity, radial velocity, angular velocity, and the sum of Hausdorff Distances). The test performance was measured using Root Mean Square Error (RMSE) and R-square (R^2^) of model predictions per dominant and non-dominant hands among the MS cohorts at the difficulty level 3. ElasticNet, SVR Radial, RF, and GBM represent respectively the Elastic net, Support Vector Regression with Radial Basis Function kernel, Random Forest, and Stochastic Gradient Boosting regression models. NA indicates cannot be computed.

| **Outcome variable** | **Models** | **Dominant hand** |  | **Non-dominant hand** |  |
| --- | --- | --- | --- | --- | --- |
|  |  | **RMSE** | **$R^2$** | **RMSE** | **$R^2$** |
| **9HPT Average (C1)** | **ElasticNet** | 0.1870 | 0.1030 | 0.2427 | 0.0366 |
|  | **SVR Radial** | 0.1201 | 0.1300 | 0.1321 | 0.0726 |
|  | **RF** | 0.1776 | 0.1247 | 0.1793 | 0.0737 |
|  | **GBM** | 0.1987 | 0.1041 | 0.1678 | 0.0971 |
| **EDSS (C4)** | **ElasticNet** | 0.1416 | 0.0302 | 0.1423 | NA |
|  | **SVR Radial** | 0.1502 | 0.0154 | 0.1484 | 0.0118 |
|  | **RF** | 0.1491 | 0.0101 | 0.1517 | 0.0039 |
|  | **GBM** | 0.1423 | 0.0318 | 0.1477 | 0.0080 |
| **CombiWISE (C5)** | **ElasticNet** | 0.1557 | 0.0352 | 0.1568 | NA |
|  | **SVR Radial** | 0.1651 | 0.0118 | 0.1594 | 0.0367 |
|  | **RF** | 0.1661 | 0.0134 | 0.1653 | 0.0113 |
|  | **GBM** | 0.1603 | 0.0297 | 0.1610 | 0.0155 |
| **NeurEx (C6)** | **ElasticNet** | 0.2722 | 0.0363 | 0.2731 | NA |
|  | **SVR Radial** | 0.2938 | 0.0162 | 0.2850 | 0.0284 |
|  | **RF** | 0.2938 | 0.0116 | 0.2893 | 0.0125 |
|  | **GBM** | 0.2811 | 0.0353 | 0.2786 | 0.0190 |

**Table S11.** The out-of-sample test performance of the clinical disability scales versus the sum of Hausdorff Distances (H.D.) using the linear regression model. The test performance was measured using Root Mean Square Error (RMSE) and R-square (R^2^) of model predictions per dominant and non-dominant hands among the MS cohorts at the difficulty level 1, 2, and 3.

| Model variables | Difficulty Levels | Dominant hand |  | Non-dominant hand |  |
| --- | --- | --- | --- | --- | --- |
| | | RMSE | $R^2$ | RMSE | $R^2$ |
| **9HPT Average (C1)**<br>**Vs.**<br>**Sum of H.D. (F24)** | **1** | 0.1879 | 0.1121 | 0.1736 | 0.0612 |
|  | **2** | 0.1707 | 0.1663 | 0.1794 | 0.0720 |
|  | **3** | 0.1769 | 0.1439 | 0.1714 | 0.1339 |
| **EDSS (C4)**<br>**Vs.**<br>**Sum of H.D. (F24)** | **1** | 0.1370 | 0.0755 | 0.1402 | 0.0306 |
|  | **2** | 0.1378 | 0.0733 | 0.1400 | 0.0364 |
|  | **3** | 0.1403 | 0.0475 | 0.1424 | 0.0429 |
| **CombiWISE (C5)**<br>**Vs.**<br>**Sum of H.D. (F24)** | **1** | 0.1484 | 0.0949 | 0.1530 | 0.0429 |
|  | **2** | 0.1491 | 0.0903 | 0.1531 | 0.0444 |
|  | **3** | 0.1541 | 0.0595 | 0.1556 | 0.0565 |
| **NeurEx (C6)**<br>**Vs.**<br>**Sum of H.D. (F24)** | **1** | 0.2622 | 0.0785 | 0.2692 | 0.0281 |
|  | **2** | 0.2633 | 0.0708 | 0.2698 | 0.0262 |
|  | **3** | 0.2714 | 0.0433 | 0.2724 | 0.0312 |

